# COVID-19 outcomes by cancer status, type, treatment, and vaccination

**DOI:** 10.1101/2022.04.19.22274047

**Authors:** Maxwell Salvatore, Miriam M. Hu, Lauren J. Beesley, Alison M. Mondul, Celeste Leigh Pearce, Christopher R. Friese, Lars G Fritsche, Bhramar Mukherjee

## Abstract

**Background:** Observational studies have identified patients with cancer as a potential subgroup of individuals at elevated risk of severe SARS-CoV-2 (COVID-19) disease and mortality. Early studies showed an increased risk of COVID-19 mortality for cancer patients, but it is not well understood how this association varies by cancer site, cancer treatment, and vaccination status.

**Methods:** Using electronic health record data from an academic medical center, we identified 259,893 individuals who were tested for or diagnosed with COVID-19 from March 10, 2020, to February 2, 2022. Of these, 41,218 tested positive for COVID-19 of whom 10,266 had a past or current cancer diagnosis. We conducted Firth-corrected, covariate-adjusted logistic regression to assess the association of cancer status, cancer type, and cancer treatment with four COVID-19 outcomes: hospitalization, intensive care unit (ICU) admission, mortality, and a composite “severe COVID-19” outcome which is the union of the first three outcomes. We examine the effect of the timing of cancer diagnosis and treatment relative to COVID diagnosis, and the effect of vaccination.

**Results:** Cancer status was associated with higher rates of severe COVID-19 infection [OR (95% CI): 1.18 (1.08, 1.29)], hospitalization [OR (95% CI): 1.18 (1.06, 1.28)], and mortality [OR (95% CI): 1.22 (1.00, 1.48)]. These associations were driven by patients whose most recent initial cancer diagnosis was within the past three years. Chemotherapy receipt was positively associated with all four COVID-19 outcomes (e.g., severe COVID [OR (95% CI): 1.96 (1.73, 2.22)], while receipt of either radiation or surgery alone were not associated with worse COVID-19 outcomes. Among cancer types, hematologic malignancies [OR (95% CI): 1.62 (1.39, 1.88)] and lung cancer [OR (95% CI): 1.81 (1.34, 2.43)] were significantly associated with higher odds of hospitalization. Hematologic malignancies were associated with ICU admission [OR (95% CI): 1.49 (1.11, 1.97)] and mortality [OR (95% CI): 1.57 (1.15, 2.11)], while melanoma and breast cancer were not associated with worse COVID-19 outcomes. Vaccinations were found to reduce the frequency of occurrence for the four COVID-19 outcomes across cancer status but those with cancer continued to have elevated risk of severe COVID [cancer OR (95% CI) among those fully vaccinated: 1.69 (1.10, 2.62)] relative to those without cancer even among vaccinated.

**Conclusion:** Our study provides insight to the relationship between cancer diagnosis, treatment, cancer type, vaccination, and COVID-19 outcomes. Our results indicate that it is plausible that specific diagnoses (e.g., hematologic malignancies, lung cancer) and treatments (e.g., chemotherapy) are associated with worse COVID-19 outcomes. Vaccines significantly reduce the risk of severe COVID-19 outcomes in individuals with cancer and those without, but cancer patients are still at higher risk of breakthrough infections and more severe COVID outcomes even after vaccination. These findings provide actionable insights for risk identification and targeted treatment and prevention strategies.

## Introduction

Coronavirus disease 2019 (COVID-19), a rapidly transmissible respiratory virus^1,2^, was first identified in Wuhan, China in November 2019^3^ and was declared a pandemic by the World Health Organization on March 11, 2020^4^. As of April 19, 2022, there have been 2,401,000 cases and 35,860 deaths in the state of Michigan^5^. Patients with cancer have been identified as a subgroup of individuals who are at high risk for severe COVID-19 disease and related mortality^2,6,7^. However, it is not well understood which cancer diagnoses or treatments may be associated with increased risk of severe outcomes related to COVID-19. Understanding how cancer diagnosis and treatment(s) are associated with COVID-19 diagnosis and outcomes is paramount since there has been a significant reduction in the supply of and demand for cancer services (e.g., urgent referrals, chemotherapy attendance) during the pandemic^8^, with research suggesting delays were to reduce cancer patient exposure to SARS-CoV-2 due to perceived increased risk for severe outcomes and to prioritize health care resources in the presence of medicine and equipment shortages^9–13^.

The evidence regarding the effect of cancer treatments (e.g., radiation, chemotherapy, and surgery) on COVID-19-related outcomes (e.g., hospitalization, ICU admission, and mortality) remains unclear. Immunosuppressive drugs are thought to increase COVID-19 severity, and some reports have stated that chemotherapy may be associated with higher risk of hospitalization or death^14,15^. However, several studies found no evidence in support of an association between chemotherapy and COVID-19 events^16–20^. There are conflicting reports on the effects of surgery^6,18,21^. Other risk factors including race, male sex, age, and comorbid conditions have been associated with COVID-19 outcomes among patients with cancer^22–24^. These studies identify potential associations that address exigent needs in cancer care during the times of COVID-19. However, these studies suffer limitations related to sample size, representativeness, early pandemic response protocols, lack of updated analysis, and replication. Few studies look at analyses by cancer type^25^, often stratifying by solid versus hematologic malignancies^21^ or limiting to one cancer type^26^. Some studies look at the timing of cancer diagnosis (e.g., diagnosed within the past year^27^, between 2010-2020^25^). Literature on COVID-19 vaccination and cancer has focused on safety^28,29^, hesitancy^30,31^, and advocacy^32^, and not on effectiveness in the population.

Building on prior literature, we use electronic health record (EHR) data from the Michigan Medicine (University of Michigan), a large, academic health care system, to quantify if having cancer was associated with increased risk of COVID-19-related hospitalization, intensive care unit (ICU) admission, 60-day all-cause mortality, and severe COVID (i.e., any of the previous outcomes). Our group has previously investigated the EHR data from Michigan Medicine for COVID-19 outcomes and risk factors in several papers, but not specifically with cancer as a focus^33–35^. In addition, we aim to determine whether cancer diagnoses, timing, types, and treatments are associated with worse COVID-19 outcomes and examine other risk factors that may increase disease severity in cancer patients contracting COVID-19. Finally, we examine emerging data on COVID-19 vaccination effect among individuals with cancer.

## Methods

### Cohort

The EHR data were collected from Michigan Medicine patients who were tested or treated for COVID-19 between the dates of March 10, 2020, through February 2, 2022. There were 351,843 individuals in this initial cohort. To distinguish between pre-existing diagnoses and diagnoses that may be related to underlying COVID-19, we restricted patient data to at least 14 days before the first COVID-19 positive test for those who tested positive, and the first COVID-19 test for those who tested negative (the “index test”). Diagnoses that are sex-specified and were discordant with the individual’s EHR-recorded sex were removed (n = 4,124). Individuals who did not have a diagnosis (cancer or otherwise) prior to the 14-day threshold were removed (n = 40,266). The analysis is further restricted to those who were adults (> 18) at the index test (n = 47,558). Finally, two individuals were removed because of their EHR-recorded age was zero or negative resulting in the analytic tested cohort of 259,893 individuals (**Figure S1**).

After exclusions, the tested cohort (n = 259,893) represents a non-probabilistic sample due to the testing protocol at Michigan Medicine, which focused initially on symptomatic and high-risk patients in the early stages of the pandemic. Of those who tested positive for COVID-19 (n = 41,218), 10,266 (24.9%) individuals had a cancer diagnosis recorded in their EHR and 4,846 (11.8%) had an initial cancer diagnosis within the past three years.

### Data

#### COVID-19 outcomes

We considered four outcomes: COVID-19-related (1) hospitalization, (2) ICU admission, and (3) mortality in addition to (4) a composite “severe COVID” outcome which is the union of (1)-(3). Those who were considered COVID-19 positive either had a positive test or were diagnosed with COVID-19. COVID-19-related hospitalization was defined as a hospitalization or discharge that occurred within 14 days prior to through 30 days after a COVID-19 diagnosis. COVID-19-related ICU admission was defined as being admitted to the ICU within 14 days prior to through 30 days after a COVID-19 diagnosis. The mortality outcome was all-cause and was defined as a death (including non-hospitalization deaths) that occurred within 14 days prior to through 60 days after a COVID-19 diagnosis (i.e., includes post-mortem COVID-19 testing). The severe COVID-19 outcome captured anyone who was identified by any of the three previous outcomes.

#### Cancer-related variables

For identifying individuals with cancer, we aggregated *International Classification of Diseases, Ninth and Tenth Revision* (ICD-9 or ICD-10) codes into broader phenotype descriptions, called phecodes (developed by Denny et al.^36^). Cancer was defined as ever having at least one phecode for cancer recorded in the patient EHRs (**Table S1**). We also define five cancer types: melanoma, hematologic malignancy, breast cancer, prostate cancer, and lung cancer (**Table S1**). Of those with cancer in the test-positive cohort, 6.4% (n = 654) were ever diagnosed with at least two of these five cancer types. We also constructed a variable corresponding to the time since the most recent initial cancer diagnosis: within the last three years, three to ten years ago, and ten or more years ago.

We considered three types of cancer treatment: chemotherapy, radiation therapy, and surgery. Chemotherapy was defined as having at least one of the chemotherapy-related Current Procedural Terminology (CPT) codes or ICD-9 or ICD-10 codes recorded in the EHR as listed in **Table S2**. Radiation therapy and surgery were defined similarly. These treatments could have taken place at any time prior to 14 days before the first positive COVID-19 test.

For the statistical analysis, we defined “radiation only” and “surgery only” variables, as radiation therapy without corresponding chemotherapy or surgery codes, and surgery without corresponding radiation or chemotherapy codes. A surgery was characterized as cancer-related only if a cancer diagnosis code was recorded during the same encounter. In addition, we constructed a variable corresponding to the time since most recent chemotherapy treatment: within last year, one to three years ago, and more than three years ago. **Figure S2** shows the distribution of cancer treatment patterns among COVID-19 positive individuals with cancer.

#### Covariates

We extracted self-reported age, sex, race/ethnicity, smoking status (never/past/current), alcohol consumption (never/past/current), and body mass index (BMI) from the EHR. Socioeconomic status measures were defined by US census tract for the year 2010 based on the patient’s residential address in the EHR and 5-year (2013-2017) American Community Survey estimates. The Neighborhood Disadvantage Index (NDI)^37^ was operationalized as quartiles and included in some covariate sets. The NDI represents the mean of: (1) proportion of female-headed families with children, (2) the proportion of households using public assistance income, (3) the proportion of people with income in the last 12 months below the poverty level, and (4) the proportion of the population (age 16 and older) unemployed. In our data, the NDI is recorded as an ordinal quartile variable using the raw mean values.

For non-cancer comorbid conditions, we used ICD-9 and ICD-10 codes from the EHR (including only those diagnoses greater than 14 days prior to first COVID-19 test or diagnosis) to construct binary disease indicators for respiratory conditions, circulatory conditions, type 2 diabetes, kidney disease, liver disease, and autoimmune disease (qualifying phecodes for each comorbidity listed in **Table S3**). The comorbidity score, which excludes cancer, was calculated as a sum of these indicators and ranges from 0-6^33^.

#### Vaccination variable

We constructed a vaccination status variable with four mutually exclusive categories: before vaccination, while partially vaccinated, while fully vaccinated (but not boosted), and after booster receipt. “Before vaccination” includes both those whose vaccination status is unvaccinated/unknown and those COVID-19 that were diagnosed prior to the availability of vaccines. Vaccination status was calculated based on the number of and manufacturer of COVID-19 vaccinations that took place at least 14 days prior to testing positive for COVID-19. Individuals who were identified as receiving a vaccine but for whom we did not have the time of the vaccination were categorized as missing. Additionally, individuals who received non-FDA-approved vaccines, including Astrazeneca, Novavax, Sinopharm, and Sinovac, were also categorized as missing. Those with missing vaccination data were excluded from vaccination analyses. In our vaccination analyses, we adjusted for a 2020 indicator that was 1 when someone was COVID-19 positive in 2020 and 0 if they were COVID-19 positive after 2020. This was to account for the lack of access to vaccinations in 2020 and for potential differences in treatments and in effects of circulating variants.

### Statistical analysis

For each of the four binary outcomes, we performed logistic regression with Firth bias correction to correct for separation issues^38^. We reported the unadjusted estimates for odds ratio (OR) as well as the estimates adjusting for three different sets of covariates (referred to as adjustment set 1, 2, and 3, respectively):

1. Age (continuous), race/ethnicity, and sex
2. Adjustment set 1 + Neighborhood Disadvantage Index (quartile)
3. Adjustment set 2 + comorbidity score

We performed analyses fitting a model of the form:

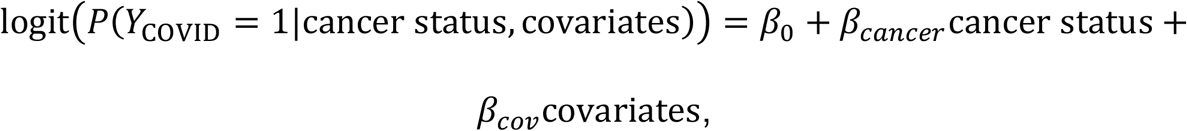

where *Y*_COVID_ is an indicator variable representing the following outcomes within the tested positive cohort comparing:

1. Those who experienced a severe COVID outcome (1) to those who did not (0)
2. Those who were hospitalized (1) to those who were not (0)
3. Those who were admitted to the ICU (1) to those who were not (0), and
4. Those who were deceased within 60 days of initial COVID-19 diagnosis (1) to those who were not (0), to assess the association between cancer status and COVID-19 outcomes.

In the above model, cancer status refers to the cancer-related indicators of interest. In addition to history of cancer, we also fit models for cancer diagnosis timing (most recent initial cancer diagnosis 0-3 years ago, 3-10 years ago, and 10 or more years ago), cancer treatment (where it represents chemotherapy, radiation only, or surgery only), chemotherapy treatment timing (most recent chemotherapy treatment less than 1 year ago, 1-3 years ago, and 3 or more years ago), and for cancer diagnosis (where is represents having a diagnosis of melanoma, a hematologic malignancy, breast cancer, prostate cancer, or lung cancer).

We also carried out interaction analyses by cancer status according to the model:

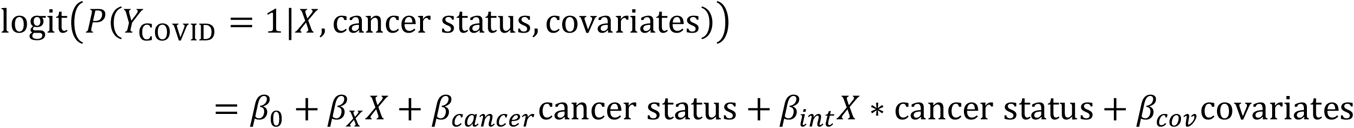

where *Y*_COVID_ is an indicator variable representing the following outcomes within the tested positive cohort comparing:

1. Severe COVID (1) to not (0)
2. Hospitalized (1) to not (0)
3. Admitted to the ICU (1) to not (0)

We did not conduct interaction analyses with the mortality outcome due to limited sample size. We considered many *X* including age (continuous and categorical), sex, BMI (continuous and categorical), race/ethnicity, alcohol consumption (binary), smoking status (never vs current and past), population density (ordinal quartile), NDI (ordinal quartile), and the comorbidity score (and each of its disease groups separately). When *X* was a component of the comorbidity score, the comorbidity score was recalculated without the disease group of interest (i.e., *X*). For the interaction analyses, we used Firth bias correction and the adjustment covariates in adjustment set 3. The p-value for the test with null hypothesis *β*_*int*_ = 0 was reported along with the p-values of subgroup associations for having cancer and not having cancer. No correction for multiple testing was applied.

Missing data were handled using complete case analysis. All analyses were performed in R statistical software version 4.1.2^39^ with Firth models using the logistf function from the logistf package^40^. The code used to conduct the analyses in this paper along with their results and figures are publicly available at: https://github.com/umich-cphds/cancer_covid.

#### Additional analyses

We conducted two additional analyses: an analysis after adjusting for vaccination status and an analysis considering the recency of the cancer diagnosis. The goal of the vaccination analysis is to assess whether vaccination status is associated with lower odds of severe COVID-19 outcomes among individuals with and without a cancer diagnosis. For this analysis, we fit a model of the form:

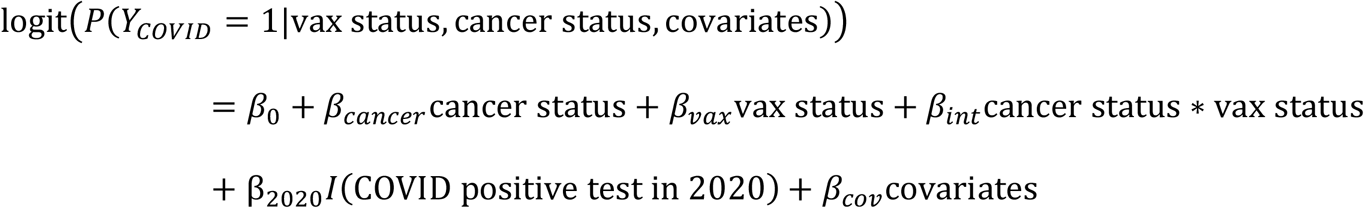

where vaccination status (described in “Covariates” subsection above) is a categorical variable with the categories before vaccination (reference), partially vaccinated, fully vaccinated (but not boosted), and boosted. A schematic representing how individuals were classified with respect to vaccination status is presented in **Figure 1B**.

**Figure 1.**
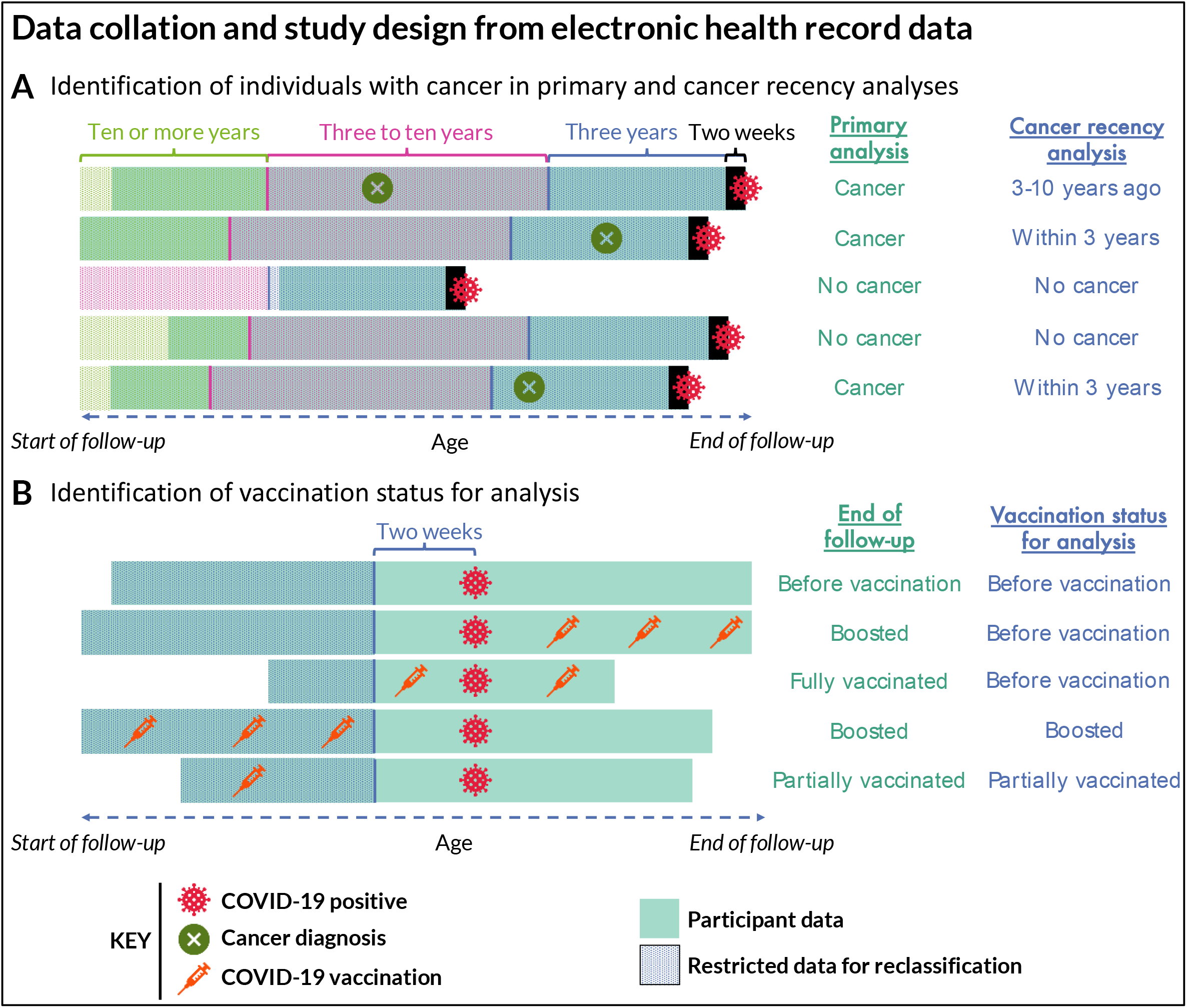
A schematic representing electronic health record scenarios and their resulting (panel A) classification of individuals with cancer for the primary and cancer recency analyses and (panel B) vaccination status for vaccine-related analyses.

In our analyses, we considered categorical variables regarding the timing of cancer diagnoses, types, and treatments. The construction of timing of cancer diagnosis (within past three years, three to ten years ago, and ten or more years ago) and chemotherapy (within past year, one to three years ago, and three or more years ago) are described above and their results reported in **Table 2** and **Table 3**, respectively. We constructed similar variables for cancer treatment considering the time of the cancer treatments relative to the positive COVID-19 test: within past three years and three or more years ago. We conducted the same analyses as presented in the “Statistical Analysis” subsection above using these variables as the independent variable (results presented in **Section S2**).

**Table 1.**
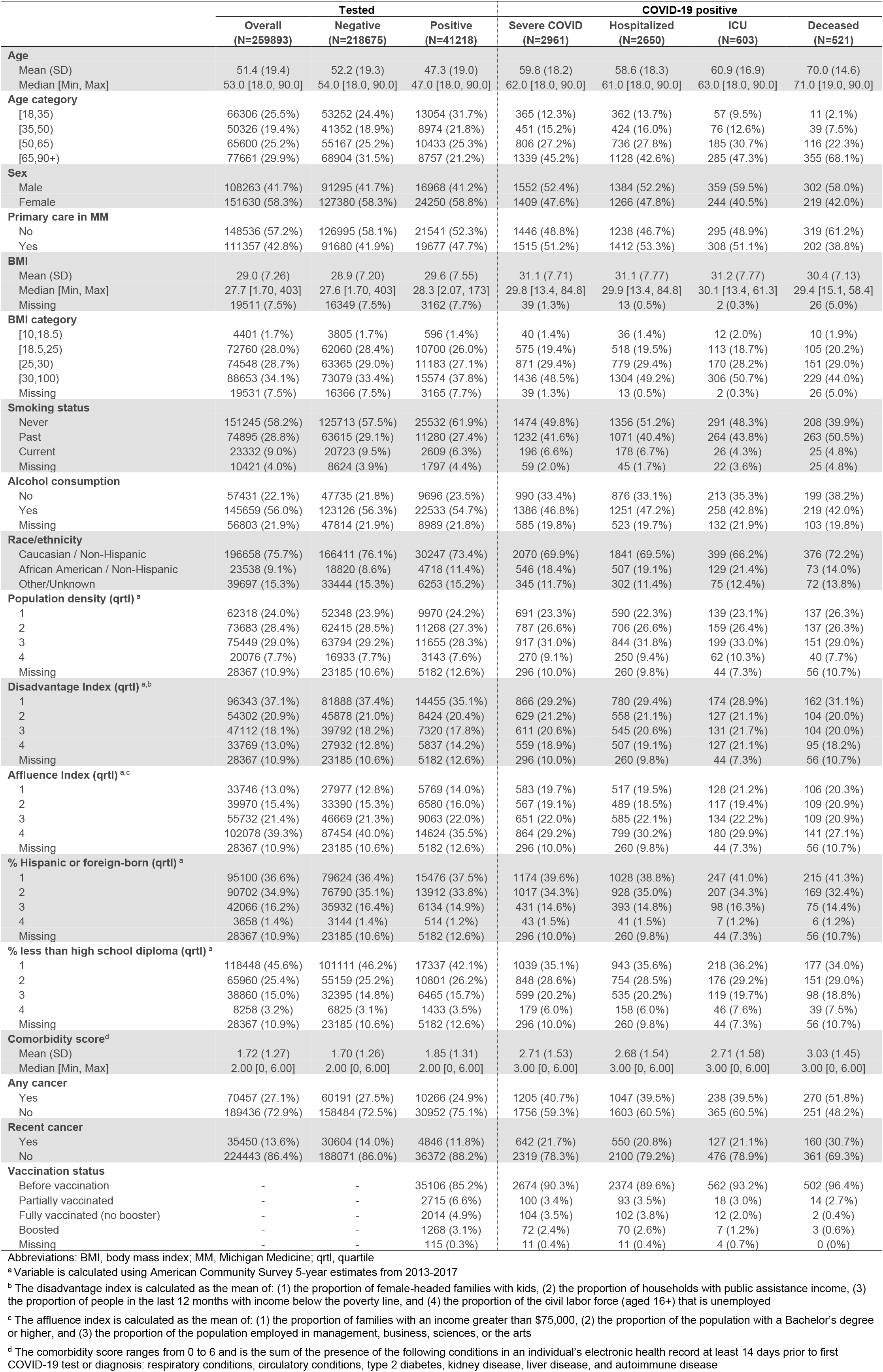
Descriptive statistics for patient characteristics among the COVID-19 tested or diagnosed cohort.

**Table 2.** Logistic regression odds ratios (95% CI) for COVID-19 outcomes by cancer status and timing of most recent initial cancer diagnosis.

|  | Outcomes |  |  |  | n |  |
| --- | --- | --- | --- | --- | --- | --- |
|  | Severe COVID | Hospitalized | ICU | Deceased | Cancer | Total |
| **Cancer (any time)** |  |  |  |  |  |  |
| Unadjusted | **2.21** (2.05, 2.39) | **2.08** (1.92, 2.26) | **1.99** (1.69, 2.35) | **3.30** (2.78, 3.93) | 10,266 | 41,218 |
| Unadjusted (Firth) | **2.21** (2.05, 2.39) | **2.08** (1.92, 2.26) | **1.99** (1.69, 2.34) | **3.30** (2.78, 3.93) | 10,266 | 41,218 |
| Adjustment 1 | **1.41** (1.29, 1.53) | **1.38** (1.27, 1.51) | **1.24** (1.04, 1.48) | **1.53** (1.28, 1.83) | 10,266 | 41,218 |
| Adjustment 2 | **1.40** (1.28, 1.52) | **1.38** (1.26, 1.51) | 1.17 (0.98, 1.41) | **1.45** (1.20, 1.76) | 9,333 | 36,036 |
| Adjustment 3 | **1.18** (1.08, 1.29) | **1.17** (1.06, 1.28) | 1.02 (0.85, 1.23) | **1.22** (1.00, 1.48) | 9,333 | 36,036 |
| **Cancer (most recent initial diagnosis within past three years)** |  |  |  |  |  |  |
| Unadjusted | **2.54** (2.31, 2.79) | **2.34** (2.12, 2.60) | **2.26** (1.84, 2.77) | **4.18** (3.42, 5.10) | 4,846 | 41,218 |
| Unadjusted (Firth) | **2.54** (2.31, 2.79) | **2.35** (2.12, 2.60) | **2.26** (1.84, 2.76) | **4.18** (3.42, 5.10) | 4,846 | 41,218 |
| Adjustment 1 | **1.57** (1.42, 1.74) | **1.52** (1.36, 1.69) | **1.35** (1.09, 1.67) | **1.86** (1.51, 2.29) | 4,846 | 41,218 |
| Adjustment 2 | **1.55** (1.39, 1.72) | **1.50** (1.34, 1.68) | **1.27** (1.01, 1.59) | **1.74** (1.39, 2.17) | 4,260 | 36,036 |
| Adjustment 3 | **1.38** (1.24, 1.54) | **1.34** (1.20, 1.51) | 1.16 (0.92, 1.45) | **1.53** (1.22, 1.92) | 4,260 | 36,036 |
| **Cancer (most recent initial diagnosis three to ten years ago)** |  |  |  |  |  |  |
| Unadjusted | **2.08** (1.87, 2.32) | **1.99** (1.78, 2.24) | **1.80** (1.41, 2.28) | **2.75** (2.15, 3.51) | 4,052 | 41,218 |
| Unadjusted (Firth) | **2.08** (1.87, 2.32) | **2.00** (1.78, 2.24) | **1.80** (1.41, 2.28) | **2.76** (2.15, 3.50) | 4,052 | 41,218 |
| Adjustment 1 | **1.29** (1.15, 1.45) | **1.30** (1.15, 1.47) | 1.11 (0.86, 1.41) | 1.20 (0.93, 1.54) | 4,052 | 41,218 |
| Adjustment 2 | **1.32** (1.17, 1.48) | **1.32** (1.16, 1.50) | 1.08 (0.83, 1.38) | 1.21 (0.93, 1.57) | 3,786 | 36,036 |
| Adjustment 3 | 1.06 (0.94, 1.20) | 1.07 (0.94, 1.21) | 0.90 (0.69, 1.16) | 0.96 (0.73, 1.25) | 3,786 | 36,036 |
| **Cancer (most recent initial diagnosis ten or more years ago)** |  |  |  |  |  |  |
| Unadjusted | **1.48** (1.21, 1.81) | **1.43** (1.16, 1.76) | **1.62** (1.09, 2.43) | **1.91** (1.22, 2.99) | 1,368 | 41,218 |
| Unadjusted (Firth) | **1.49** (1.21, 1.81) | **1.43** (1.16, 1.76) | **1.65** (1.08, 2.41) | **1.95** (1.21, 2.96) | 1,368 | 41,218 |
| Adjustment 1 | 1.15 (0.93, 1.40) | 1.14 (0.91, 1.40) | 1.30 (0.85, 1.91) | 1.30 (0.80, 1.99) | 1,368 | 41,218 |
| Adjustment 2 | 1.12 (0.90, 1.38) | 1.11 (0.88, 1.38) | 1.14 (0.72, 1.73) | 1.14 (0.67, 1.81) | 1,287 | 36,036 |
| Adjustment 3 | 0.90 (0.72, 1.11) | 0.90 (0.71, 1.12) | 0.96 (0.60, 1.45) | 0.90 (0.53, 1.44) | 1,287 | 36,036 |

Adjustment 1: Age, race/ethnicity, sex;
Adjustment 2: Adjustment 1 + Disadvantage Index (quartile)
Adjustment 3: Adjustment 2 + comorbidity score.
Bolded point estimates represents statistical significance at the 95% confidence level.

**Table 3.** Logistic regression odds ratios (95% CI) for COVID-19 outcomes by cancer treatment and timing of most recent chemotherapy.

|  | Outcomes |  |  |  | n |  |
| --- | --- | --- | --- | --- | --- | --- |
|  | Severe COVID | Hospitalized | ICU | Deceased | Treatment group | Total |
| **Chemotherapy (any time)** |  |  |  |  |  |  |
| Unadjusted | **3.92** (3.50, 4.38) | **3.75** (3.33, 4.21) | **3.16** (2.50, 4.00) | **5.04** (3.98, 6.40) | 2,449 | 41,218 |
| Unadjusted (Firth) | **3.92** (3.50, 4.38) | **3.75** (3.33, 4.21) | **3.17** (2.49, 3.99) | **5.06** (3.97, 6.39) | 2,449 | 41,218 |
| Adjustment 1 | **2.56** (2.28, 2.88) | **2.54** (2.25, 2.87) | **1.99** (1.55, 2.53) | **2.46** (1.92, 3.13) | 2,449 | 41,218 |
| Adjustment 2 | **2.58** (2.28, 2.91) | **2.58** (2.27, 2.92) | **1.89** (1.46, 2.42) | **2.32** (1.78, 3.00) | 2,217 | 36,036 |
| Adjustment 3 | **1.96** (1.73, 2.22) | **1.96** (1.72, 2.24) | **1.50** (1.15, 1.94) | **1.73** (1.32, 2.25) | 2,217 | 36,036 |
| **Most recent chemotherapy treatment within past year** |  |  |  |  |  |  |
| Unadjusted | **4.80** (4.08, 5.63) | **4.53** (3.83, 5.36) | **3.64** (2.60, 5.09) | **6.60** (4.81, 9.05) | 938 | 41,218 |
| Unadjusted (Firth) | **4.80** (4.08, 5.63) | **4.54** (3.83, 5.35) | **3.67** (2.59, 5.07) | **6.65** (4.80, 9.02) | 938 | 41,218 |
| Adjustment 1 | **3.18** (2.69, 3.76) | **3.11** (2.60, 3.69) | **2.29** (1.60, 3.18) | **3.29** (2.36, 4.51) | 938 | 41,218 |
| Adjustment 2 | **3.27** (2.73, 3.90) | **3.22** (2.68, 3.86) | **2.25** (1.55, 3.17) | **2.99** (2.07, 4.20) | 818 | 36,036 |
| Adjustment 3 | **2.70** (2.25, 3.23) | **2.67** (2.21, 3.20) | **1.91** (1.31, 2.70) | **2.42** (1.67, 3.42) | 818 | 36,036 |
| **Most recent chemotherapy treatment one to three years ago** |  |  |  |  |  |  |
| Unadjusted | **3.70** (2.91, 4.70) | **3.39** (2.63, 4.38) | **3.17** (1.93, 5.19) | **5.19** (3.22, 8.35) | 467 | 41,218 |
| Unadjusted (Firth) | **3.72** (2.91, 4.70) | **3.41** (2.63, 4.37) | **3.25** (1.92, 5.14) | **5.31** (3.21, 8.27) | 467 | 41,218 |
| Adjustment 1 | **2.40** (1.86, 3.06) | **2.28** (1.75, 2.94) | **2.02** (1.19, 3.23) | **2.61** (1.57, 4.12) | 467 | 41,218 |
| Adjustment 2 | **2.47** (1.90, 3.18) | **2.39** (1.81, 3.12) | **1.89** (1.07, 3.09) | **2.56** (1.49, 4.14) | 422 | 36,036 |
| Adjustment 3 | **1.92** (1.47, 2.48) | **1.86** (1.41, 2.43) | 1.53 (0.86, 2.50) | **1.97** (1.14, 3.19) | 422 | 36,036 |
| **Most recent chemotherapy treatment more than three years ago** |  |  |  |  |  |  |
| Unadjusted | **3.28** (2.77, 3.89) | **3.24** (2.71, 3.87) | **2.74** (1.91, 3.93) | **3.62** (2.47, 5.31) | 1,044 | 41,218 |
| Unadjusted (Firth) | **3.29** (2.76, 3.89) | **3.25** (2.71, 3.86) | **2.77** (1.90, 3.91) | **3.67** (2.46, 5.28) | 1,044 | 41,218 |
| Adjustment 1 | **2.13** (1.78, 2.53) | **2.19** (1.82, 2.62) | **1.76** (1.20, 2.50) | **1.74** (1.15, 2.52) | 1,044 | 41,218 |
| Adjustment 2 | **2.10** (1.74, 2.52) | **2.16** (1.78, 2.60) | **1.62** (1.08, 2.34) | **1.73** (1.13, 2.55) | 977 | 36,036 |
| Adjustment 3 | **1.46** (1.21, 1.76) | **1.51** (1.24, 1.83) | 1.21 (0.80, 1.76) | 1.18 (0.77, 1.75) | 977 | 36,036 |
| **Radiation only** |  |  |  |  |  |  |
| Unadjusted | **2.42** (1.79, 3.26) | **2.24** (1.63, 3.09) | **2.41** (1.31, 4.42) | **4.17** (2.37, 7.35) | 394 | 41,218 |
| Unadjusted (Firth) | **2.44** (1.79, 3.25) | **2.27** (1.62, 3.08) | **2.51** (1.31, 4.34) | **4.32** (2.36, 7.25) | 394 | 41,218 |
| Adjustment 1 | 1.27 (0.93, 1.71) | 1.26 (0.90, 1.73) | 1.27 (0.66, 2.22) | 1.52 (0.82, 2.58) | 394 | 41,218 |
| Adjustment 2 | 1.11 (0.78, 1.55) | 1.12 (0.77, 1.59) | 1.01 (0.46, 1.90) | 1.20 (0.57, 2.21) | 353 | 36,036 |
| Adjustment 3 | 0.97 (0.68, 1.36) | 0.98 (0.67, 1.39) | 0.90 (0.41, 1.70) | 0.99 (0.47, 1.84) | 353 | 36,036 |
| **Surgery only** |  |  |  |  |  |  |
| Unadjusted | **2.02** (1.10, 3.68) | **2.01** (1.08, 3.76) | 1.54 (0.38, 6.25) | 1.11 (0.15, 8.00) | 111 | 41,218 |
| Unadjusted (Firth) | **2.09** (1.10, 3.62) | **2.09** (1.07, 3.70) | 1.91 (0.40, 5.49) | 1.66 (0.19, 6.02) | 111 | 41,218 |
| Adjustment 1 | 1.21 (0.63, 2.13) | 1.27 (0.65, 2.28) | 1.11 (0.23, 3.22) | 0.70 (0.08, 2.59) | 111 | 41,218 |
| Adjustment 2 | 1.23 (0.62, 2.21) | 1.28 (0.63, 2.34) | 1.16 (0.24, 3.38) | 0.77 (0.09, 2.88) | 102 | 36,036 |
| Adjustment 3 | 0.93 (0.47, 1.69) | 0.98 (0.48, 1.81) | 0.92 (0.19, 2.70) | 0.54 (0.06, 2.05) | 102 | 36,036 |

Adjustment 1: Age, race/ethnicity, sex
Adjustment 2: Adjustment 1 + Disadvantage Index (quartile)
Adjustment 3: Adjustment 2 + comorbidity score.
Bolded point estimates represents statistical significance at the 95% confidence level.

## Results

The analytic cohort of COVID-19 tested individuals (n = 259,893) was 41.7% (n = 108,263) male, had a median (interquartile range [IQR]) age of 53.0 (34.0, 67.0) years, a mean (standard deviation [SD]) BMI of 29.0 (7.3) kg/m^2^, and a mean (SD) comorbidity score of 1.7 (1.3). Of those who were tested, 15.9% (n = 41,218) were positive for COVID-19. The COVID-19 positive cohort was slightly younger (median [IQR] age: 47.0 [30.0, 62.0] years) and had a higher comorbidity score (mean [SD] 1.9 [1.3]). In test-positive cohort, as COVID-19 outcome severity increased, age, proportion male, and comorbidity score tended to increase. The descriptive statistics from this cohort are presented in **Table 1**.

In the test-positive cohort, 24.9% (n = 10,266) had a prior diagnosis of cancer. Among all individuals who tested positive for COVID-19 in our data, patients with cancer comprised 40.7% (n = 1,205) of those with severe COVID, 39.5% (n = 1,047) of those who were hospitalized, 39.5% (n = 238) of those who were admitted to the ICU, and 51.8% (n = 270) of those who died. The number and proportion of individuals with and without cancer is presented in **Figure 2**.

**Figure 2.**
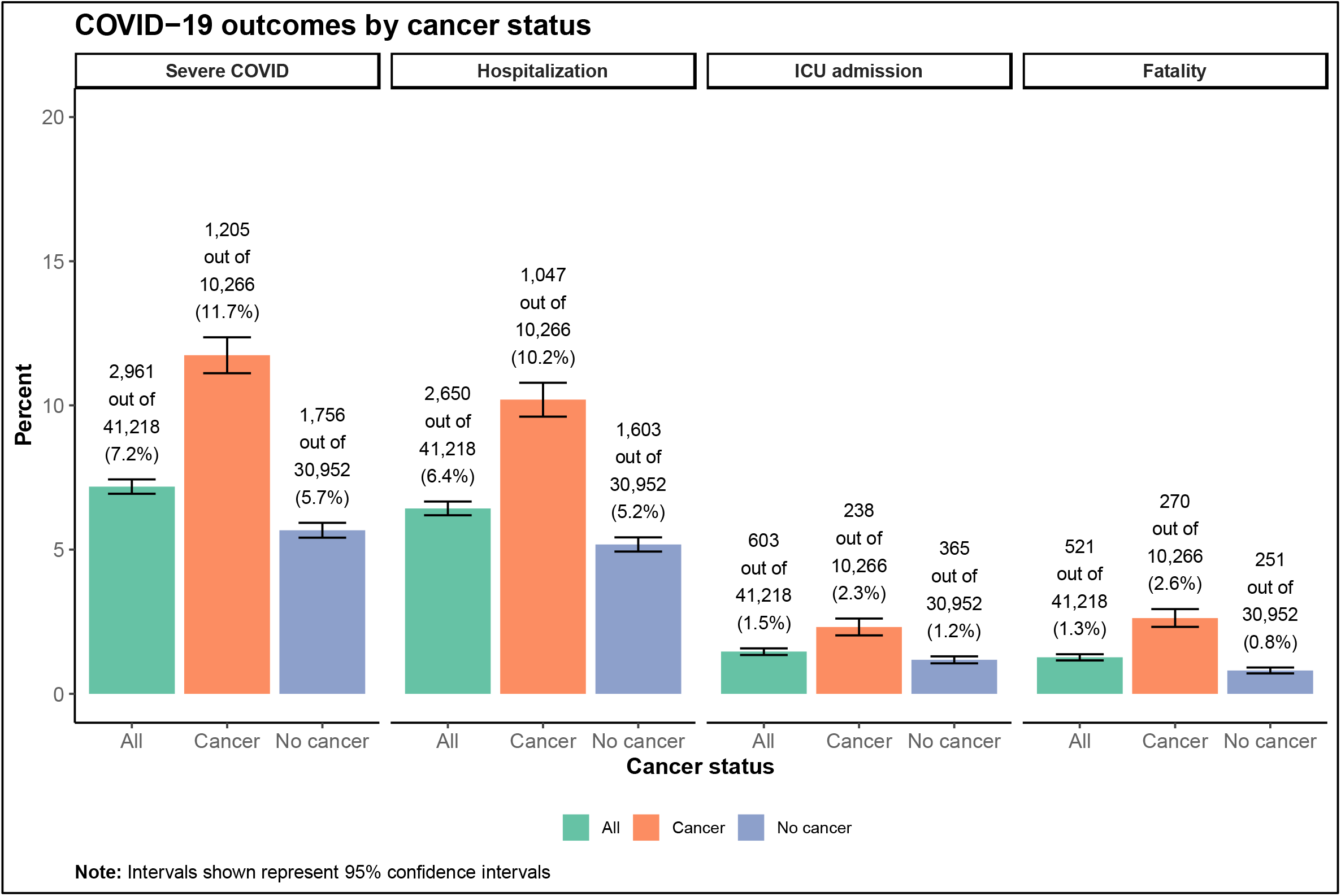
COVID-19 outcomes by cancer status. The bars present raw proportion of individuals with the outcome (*Y* = 1) overall (green), among those with cancer (orange), and among those without cancer (purple). The first panel (“Severe COVID”) represents the proportion of individuals who were hospitalized, admitted to the ICU, or died because of COVID-19. The error bars represent the 95% confidence intervals.

Among those who tested positive for COVID-19, having a cancer diagnosis was significantly associated with higher rates of severe COVID [OR (95% CI): 1.18 (1.08, 1.29)], hospitalization [OR (95% CI): 1.17 (1.06, 1.28)], and 60-day all-cause mortality [OR (95% CI): 1.22 (1.00, 1.48)]. We see these associations are driven by people whose most recent initial cancer diagnosis was within the past three years (severe COVID [OR (95% CI): 1.38 (1.24, 1.54)], hospitalization [OR (95% CI): 1.34 (1.20, 1.51)], and 60-day all-cause mortality [OR (95% CI): 1.53 (1.22, 1.92)]. Neither a history of cancer [OR (95% CI): 1.02 (0.85, 1.23)] nor an initial cancer diagnosis within the past three years [OR (95% CI): 1.16 (0.92, 1.45)] was associated with an increased rate of ICU admission. After adjustment, there was not association between most recent initial cancer diagnoses three or more years ago and any of the COVID-19 outcomes. Our results for cancer status and COVID-19 outcomes are shown in **Table 2**.

Our analyses by cancer treatment found a significant association between chemotherapy receipt and severe COVID [OR (95% CI): 1.96 (1.73, 2.22)], hospitalization [OR (95% CI): 1.96 (1.72, 2.24)], ICU admission [OR (95% CI): 1.50 (1.15, 1.94)], and 60-day all-cause mortality [OR (95% CI): 1.73 (1.32, 2.25)]. Most recent chemotherapy within the past year was most strongly associated with severe COVID [OR (95% CI): 2.70 (2.25, 3.23)], with the association weakening as time since last chemotherapy treatment increased (one to three years ago [OR (95% CI): 1.92 (1.47, 2.48)]; more than three years ago [OR (95% CI): 1.46 (1.21, 1.76)]). After covariate adjustment, surgery-only and radiation-only were not associated with any COVID-19 outcome. These results are presented in **Table 3**. Chemotherapy treatment (at any time) remained associated with all four outcomes after exclusion of patients with hematologic malignancies (e.g., severe COVID OR (95% CI): 1.76 (1.51, 2.05); **Table S4**). When we examined treatments administered within three years of testing positive for COVID-19, radiation-only was associated with severe COVID [OR (95% CI): 1.74 (1.14, 2.59)], hospitalization [OR (95% CI): 1.86 (1.20, 2.79)], and ICU admission [OR (95% CI): 2.77 (1.41, 4.93)] (**Table S5**).

In our fully adjusted analyses by cancer type, we found that those with hematologic malignancies [OR (95% CI): 1.62 (1.39, 1.88)] and lung cancer [OR (95% CI): 1.81 (1.34, 2.43)] had significantly higher rates of hospitalization. Additionally, hematologic malignancy diagnoses were significantly associated with higher risk of severe COVID [OR (95% CI): 1.66 (1.43, 1.91)], ICU admission [OR (95% CI): 1.49 (1.11, 1.97)], and mortality [OR (95% CI): 1.57 (1.15, 2.11)]. Melanoma and breast cancer diagnoses did not show a significant difference in any of the outcomes in the fully adjusted model, though the results suggest potential for melanoma (e.g., hospitalization OR (95% CI): 1.18 (0.99, 1.41)). Prostate cancer was negatively associated with hospitalization and ICU admission. Results from our analyses of COVID-19 outcomes by cancer type are shown in **Table 4**.

**Table 4.**
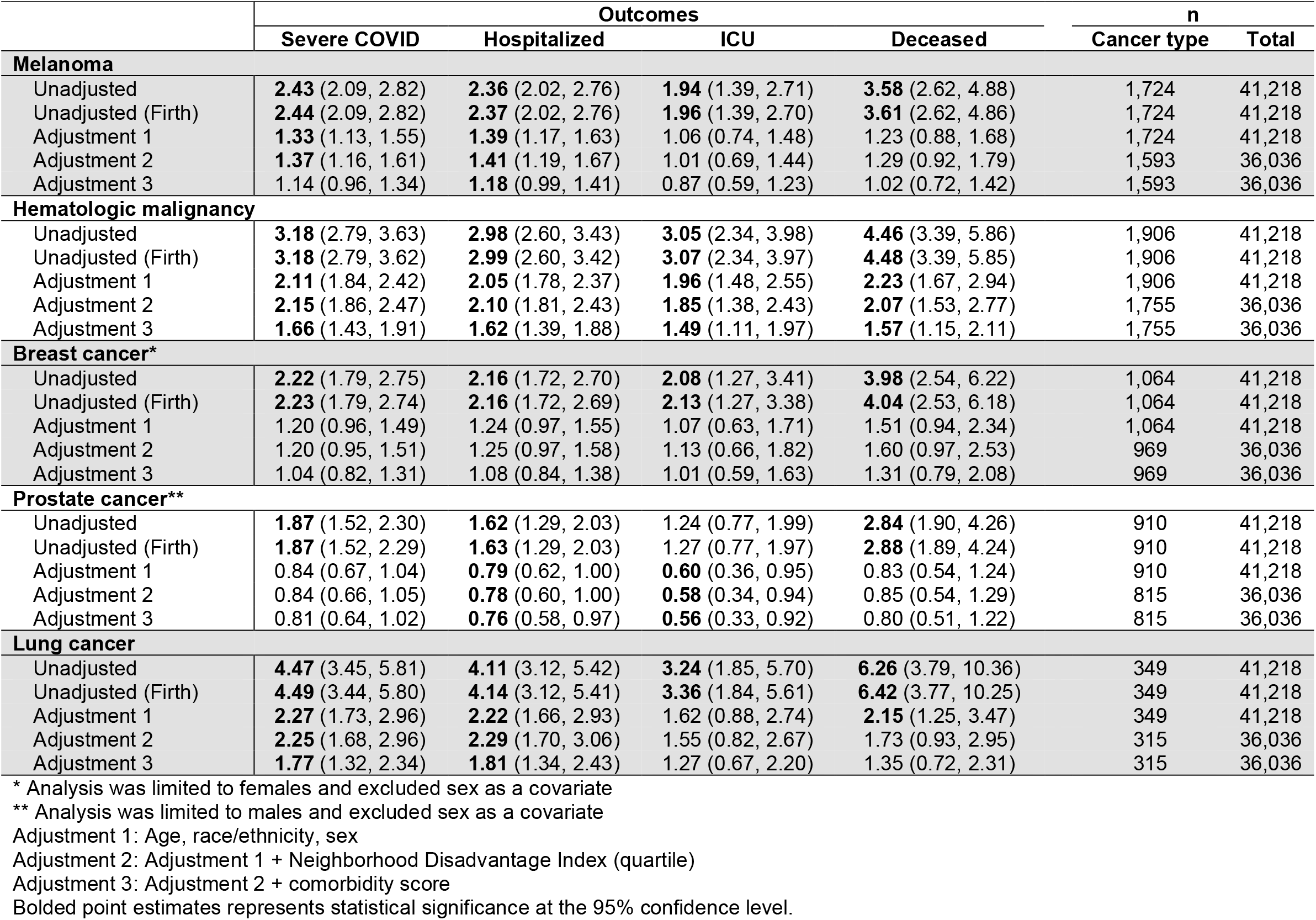
Logistic regression odds ratios (95% CI) for COVID-19 outcomes by cancer type.

**Table 5.** Vaccination status odds ratios (95% CI) for COVID-19 outcomes by cancer status strata and p-value for vaccination status-cancer status interaction

| Outcome | Cancer Status |  | P-values |  |  | n |  |  |
| --- | --- | --- | --- | --- | --- | --- | --- | --- |
|  | Cancer<br>OR (95% CI) | No cancer<br>OR (95% CI) | Cancer | No cancer | Interaction | Outcome and<br>vaccine | Vaccine | Total |
| **Severe COVID** |  |  |  |  |  |  |  |  |
| Partially vaccinated | 0.75 (0.50, 1.10) | **0.57** (0.40, 0.80) | 0.15 | <0.01 | 0.32 | 64 | 1,102 | 35,940 |
| Fully vaccinated (no booster) | **0.61** (0.45, 0.82) | **0.43** (0.30, 0.59) | <0.01 | <0.01 | 0.11 | 94 | 1,797 | 35,940 |
| Boosted | **0.35** (0.25, 0.46) | **0.29** (0.20, 0.40) | <0.01 | <0.01 | 0.45 | 87 | 2,360 | 35,940 |
| **Hospitalization** |  |  |  |  |  |  |  |  |
| Partially vaccinated | 0.88 (0.58, 1.29) | **0.63** (0.43, 0.89) | 0.52 | <0.01 | 0.23 | 62 | 1,102 | 35,940 |
| Fully vaccinated (no booster) | **0.73** (0.53, 0.97) | **0.49** (0.35, 0.68) | 0.03 | <0.01 | 0.08 | 92 | 1,797 | 35,940 |
| Boosted | **0.37** (0.27, 0.50) | **0.33** (0.23, 0.46) | <0.01 | <0.01 | 0.62 | 81 | 2,360 | 35,940 |
| **ICU admission** |  |  |  |  |  |  |  |  |
| Partially vaccinated | 0.35 (0.07, 1.02) | **0.42** (0.14, 0.94) | 0.06 | 0.03 | 0.84 | 6 | 1,102 | 35,940 |
| Fully vaccinated (no booster) | 0.66 (0.32, 1.21) | **0.16** (0.03, 0.45) | 0.19 | <0.01 | 0.03 | 11 | 1,797 | 35,940 |
| Boosted | **0.41** (0.20, 0.75) | **0.38** (0.16, 0.73) | <0.01 | <0.01 | 0.87 | 16 | 2,360 | 35,940 |
| **Fatality** |  |  |  |  |  |  |  |  |
| Partially vaccinated | **0.24** (0.05, 0.70) | **0.16** (0.02, 0.59) | <0.01 | <0.01 | 0.70 | 3 | 1,102 | 35,940 |
| Fully vaccinated (no booster) | **0.10** (0.02, 0.29) | **0.04** (0.00, 0.25) | <0.01 | <0.01 | 0.45 | 2 | 1,797 | 35,940 |
| Boosted | **0.29** (0.15, 0.52) | **0.14** (0.03, 0.39) | <0.01 | <0.01 | 0.25 | 12 | 2,360 | 35,940 |

Models were adjusted for age, race/ethnicity, sex, Disadvantage Index (quartile) and comorbidity score, and an indicator variable for whether COVID-19 was diagnosed in 2020 (1) or not (0). Fully vaccinated does not include individuals who received a booster.
Bolded point estimates represents statistical significance at the 95% confidence level.

Results from our interaction analyses for cancer (diagnosed at any time) and severe COVID (**Figure S3**), hospitalization (**Figure S4**), and ICU admission (**Figure S5**) are presented in the **Supplementary Section 1.1**. Interaction analyses for recent cancer diagnosis (within 3 years of COVID-19 positive test) and severe COVID (**Figure S6**), hospitalization (**Figure S7**), and ICU admission (**Figure S8**) are presented in **Supplementary Section 2**. 60-day all-cause mortality was not considered as an outcome for interaction analyses due to sample size.

### Vaccination analysis results

The vaccination results demonstrate that increased vaccination coverage – from partially vaccinated (OR [95% CI]: 0.75 [0.50, 1.10] among those with cancer; 0.57 [0.40, 0.80] among those without) to fully vaccinated (OR [95% CI]: 0.61 [0.45, 0.82] among those with cancer; 0.43 [0.30, 0.59] among those without) to boosted (OR [95% CI]: 0.35 [0.25, 0.46] among those with cancer; 0.29 [0.20, 0.40] among those without) - is associated with significantly lowers odds of severe COVID-19. This trend is seen across hospitalization, ICU admission, and fatality. Moreover, this effect is similarly effective among both those with and without cancer (i.e., no evidence of interaction). The results are summarized in **Table 4**. However, across every stratum of vaccination status, individuals with cancer are at elevated risk for severe COVID (e.g., cancer OR (95% CI) for severe COVID among fully vaccinated individuals: 1.69 (1.10, 2.62); **Figure 3**).

**Figure 3.**
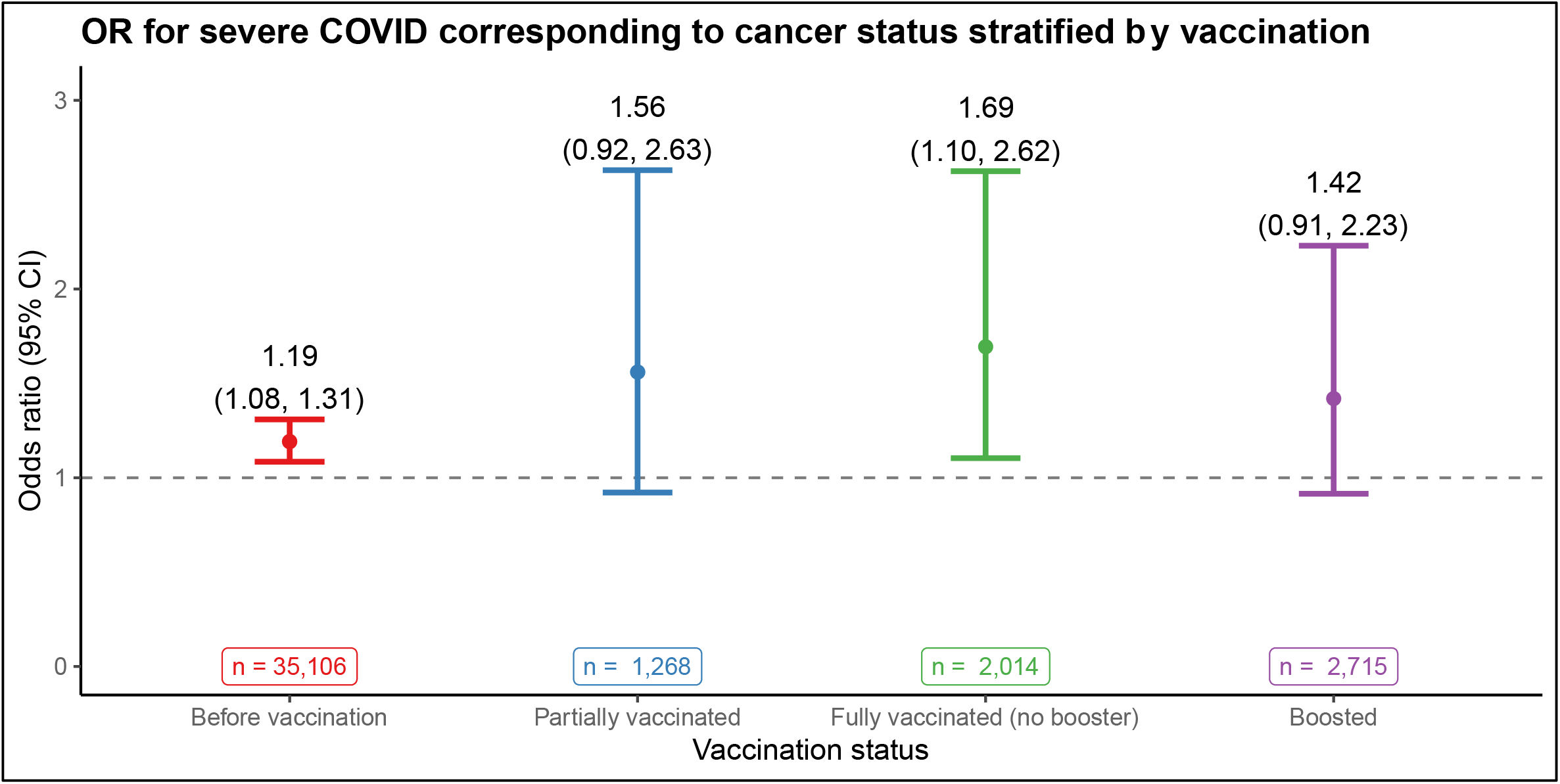
Odds ratio for severe COVID-19 corresponding to cancer status by COVID-19 vaccination strata. “Before vaccination” includes those whose vaccination status is unvaccinated/unknown as well as those who were diagnosed with COVID-19 prior to the availability of COVID-19 vaccines. Additionally, an indicator variable indicating whether the COVID-19 diagnosis was made in 2020 (1) or not (0) was added as a covariate in the model to account for differences in strains.

## Discussion

In this study, we examined a cohort of 41,218 individuals who tested positive for COVID-19 among 259,893 tested patients to determine if cancer diagnosis is associated with increased COVID-19 susceptibility or outcome severity (hospitalization, ICU admission, mortality). Consistent with Sun et al.^41^, we found positive associations between cancer status and the four severity outcomes: severe COVID [OR (95% CI): 1.18 (1.08, 1.29)], hospitalization [OR (95% CI): 1.17 (1.06, 1.28)], and 60-day all-cause mortality [OR (95% CI): 1.22 (1.00, 1.48)]. We find that these associations are driven by individuals whose most recent initial cancer diagnosis was within the past three years since there were no associations found in those whose most recent initial cancer diagnosis was more than three years ago.

We also examined differences in risk by cancer treatment and by cancer type. Our study provides evidence that chemotherapy receipt is associated with worse COVID-19 outcomes, including hospitalization [OR (95% CI): 1.96 (1.72, 2.24)]. Moreover, this association exhibited attenuation as time since most recent chemotherapy treatment increased and remained significant when three or more years ago. While there are some smaller studies found no difference in outcomes for chemotherapy patients^16^, a recent paper by Chavez-MacGregor et al.^42^ found that recent chemotherapy treatment is associated with mortality in COVID-19 patients. This finding is consistent with both chemotherapy’s potential for immunosuppression^43^ and our finding that hematologic malignancy diagnoses were associated with higher odds of hospitalization, for which myelosuppressive chemotherapy is a primary form of treatment^44^. We also found that chemotherapy receipt continued to be positively associated with higher rates of severe COVID [OR (95% CI): 1.76 (1.51, 2.05)], hospitalization [OR (95% CI): 1.76 (1.50, 2.07)], and 60-day all-cause mortality [OR (95% CI): 1.66 (1.20, 2.26)] even when the analysis excluded hematologic malignancies (**Table S4**). While we found these associations plausible, it is likely that they are overstated since chemotherapy could be a proxy for patients with high-stage cancer. Radiation-only was not found to be significantly associated with COVID-19 ICU admission or mortality, though our sensitivity analysis suggests recent radiation-only treatment may be associated with severe COVID, COVID-19 hospitalization, and ICU admission (**Table S5**). Given this information, extra COVID-19 protection measures, increased surveillance, and early and aggressive COVID-19 treatment may be warranted for patients who receive chemotherapy.

We identified that hematologic malignancies and lung cancer diagnoses were associated with higher odds of severe COVID and COVID-19 hospitalization. Hematologic malignancies were additionally associated with higher likelihood of ICU admission and mortality (a result reported by Fu et al.^45^). However, melanoma, breast cancer, and prostate cancer were not. In fact, we found that prostate cancer was associated with lower odds of hospitalization and ICU admission. We believe this finding is attributable to high rates of prostate cancer diagnosis among individuals with indolent disease who are otherwise healthy. Our cancer type analysis suggests that only hematologic malignancies are associated with COVID-19 mortality (**Table 4**). This result contrasts with those from Fillmore et al.^25^, which concludes there is excess mortality among COVID-19 positive patients with melanoma, breast, and prostate cancer diagnoses. Other studies, like Vuagnat et al.’s^46^ breast cancer cohort, conclude, in alignment with our results, that COVID-19 mortality appears to depend more on comorbid conditions than on cancer treatments.

In earlier studies on COVID-19 and cancer, sample sizes for individuals with cancer and concurrent COVID-19 were in the hundreds for most studies, resulting in small numbers of events^47^. Thus, results showing large treatment effects in a small number of patients should be interpreted cautiously^47^. Some studies focused on specific populations such as the US Veterans Affairs Healthcare System^25^ and may not be representative of the general population. Moreover, many of these studies were conducted earlier in the pandemic, when testing was targeted toward symptomatic cases, potentially inducing bias due to a lack of representativeness of individuals who were officially diagnosed with COVID-19. More recent studies have been conducted with cancer-only cohorts^48,49^, including one with almost 5,000 cases^50^, and often focus only on COVID-19-specific mortality. Regarding cancer patients and vaccines, research has focused on immunological response and safety^28,29^, vaccine hesitancy^30,31^, and tend to advocate for COVID-19 vaccination for cancer patients while urging for more research on this area^32^.

Our study has important limitations. First, as it is reliant on Michigan Medicine EHR data, the quality of the data depends on patient utilization of Michigan Medicine services. We estimate that only 48.9% of all COVID-19 positive patients received primary care services at Michigan Medicine. As such, many patients may have been hospitalized elsewhere and not had their downstream COVID-19 test results or outcomes captured outside of the Michigan Medicine EHR. Second, COVID-19 testing is not a simple random process. Though it has since expanded from targeted protocols at the start of the pandemic, factors like health insurance and access to and utilization of care present challenges to obtaining a representative testing cohort. Third, we define individuals with cancer as the presence of a cancer related phecode *at any point* in their Michigan Medicine EHR. As a result, there are concerns about survival bias – that cancer patients represent individuals who are cancer survivors, and these individuals are likely to differ systematically from current cancer patients. However, of the 10,266 COVID-19 positive individuals with a cancer diagnosis, 47% (n = 4,846) had an initial cancer diagnosis in the three years prior to testing positive. We also conducted additional analyses by timing of most recent cancer diagnosis (within 3 years, 3-10 years, 10 or more years). Fourth, the data comes from a single site - specifically a large, academic healthcare system in Michigan – and may not be representative of the state or US population, limiting the generalizability of the results. Fifth, the outcomes are defined using time periods around the index test, which means the outcomes could be the result of something unrelated to COVID-19 (e.g., death due to end-stage cancer rather than COVID-19). We calculated the absolute rates of hospitalization, ICU admission, and fatality in COVID-19 test-positive and in matched and unmatched test-negative patients (**Table 6**). We see that the absolute rates of ICU admission and fatality were higher in the COVID-19 positive cohort than in the COVID-19 negative cohorts. Additionally, the cancer-no cancer outcome rate ratios are stable across COVID-19 positive and negative status, suggesting that there is not a synergistic effect between COVID-19 and cancer for these outcomes.

**Table 6.** Rate of hospital stays, critical care stays, and deaths in COVID-19 positives, unmatched test-negatives, and age- and sex-matched test-negatives.

| *Time window relative to index test (days)* | **n (%)** |  |  |  |  |
| --- | --- | --- | --- | --- | --- |
|  | **Overall** | **No Hospital Stay** | **Hospital Inpatient Care\*,\*\*** | **Critical Care\*\*** | **Death** |
|  |  | *(-14, +30)* | *(-14, +30)* | *(-14, +30)* | *(-14, +60)* |
| Total | 304,546 | 274,395 (90.1) | 30,151 (9.9) | 7,106 (2.3) | 3,024 (1.0) |
| **COVID-19 positive** (n total) | 47,424 | 42,763 (90.2) | 4,661 (9.8) | 1,425 (3.0) | 651 (1.4) |
| Cancer | 10,266 | 8,868 (86.4) | 1,398 (13.6) | 362 (3.5) | 270 (2.6) |
| No cancer | 37,158 | 33,895 (91.2) | 3,263 (8.8) | 1,063 (2.9) | 381 (1.0) |
| **COVID-19 negative (unmatched)** (n total) | 257,122 | 231,632 (90.1) | 25,490 (9.9) | 5,681 (2.2) | 2,373 (0.9) |
| Cancer | 60,193 | 52,497 (87.2) | 7,696 (12.8) | 1,607 (2.7) | 1,091 (1.8) |
| No cancer | 196,929 | 179,135 (91.0) | 17,794 (9.0) | 4,074 (2.1) | 1,282 (0.7) |
| **COVID-19 negative (age and sex matched 1:5)** (n total) | 237,105 | 214,559 (90.5) | 22,546 (9.5) | 4,945 (2.1) | 1,923 (0.8) |
| Cancer | 51,508 | 45,074 (87.5) | 6,434 (12.5) | 1,350 (2.6) | 866 (1.7) |
| No cancer | 185,597 | 169,485 (91.3) | 16,112 (8.7) | 3,595 (1.9) | 1,057 (0.6) |

The index test is the first positive test among COVID-19 positive individuals and the first COVID-19 test among test-negative individuals
\* CPT4 Procedure codes: 99221-99239, 99460-99462, or 99477
\*\* CPT4 Procedure codes: 99289-99298, or 99466-99476

Our study contributes important information to the area of cancer and treatment in the time of COVID-19. Specifically, cancer status alone appears to be associated with higher rates of COVID-19-related hospitalization, ICU admission, and mortality. This association is driven be people with recent cancer diagnoses. The existence and strength of an association is different based on cancer diagnosis (e.g., hematologic malignancies were associated with worse COVID-19 outcomes while breast cancer was not) and treatment (e.g., chemotherapy was associated with worse COVID-19 outcomes while surgery only was not). Additionally, chemotherapy appeared to be associated with worse COVID-19 outcomes even after the exclusion of cancer patients with hematologic malignancies. Finally, we provide evidence that vaccination is effective in reducing severe COVID in cancer patients. Future research should consider post-acute sequalae of COVID-19 (PASC, or “long COVID”) as an outcome and look more closely at the role cancer types, treatments, and COVID-19 vaccination play in COVID-19 outcomes.

## Data Availability

The individual level data cannot be shared publicly due to patient confidentiality. The data underlying the results presented in the study are available from University of Michigan Data Office for Clinical & Translational Research for researchers who meet the criteria for access to confidential data.

## Institutional Review Board statement

The University of Michigan Medical School institutional review board reviewed the study and determined that it is exempt. Ethical review and approval were waived for this study per institutional policy.

