## Supplementary Material for "COVID-19 outcomes by cancer status, type, treatment, and vaccination" (from another version of this preprint: version 1, 2022-04-21)

**Section 1**. Tables and figures accompanying primary and vaccination results.

**Figure S1**. Flowchart describing derivation of COVID-19 tested analytic cohort from raw COVID-19 tested data.

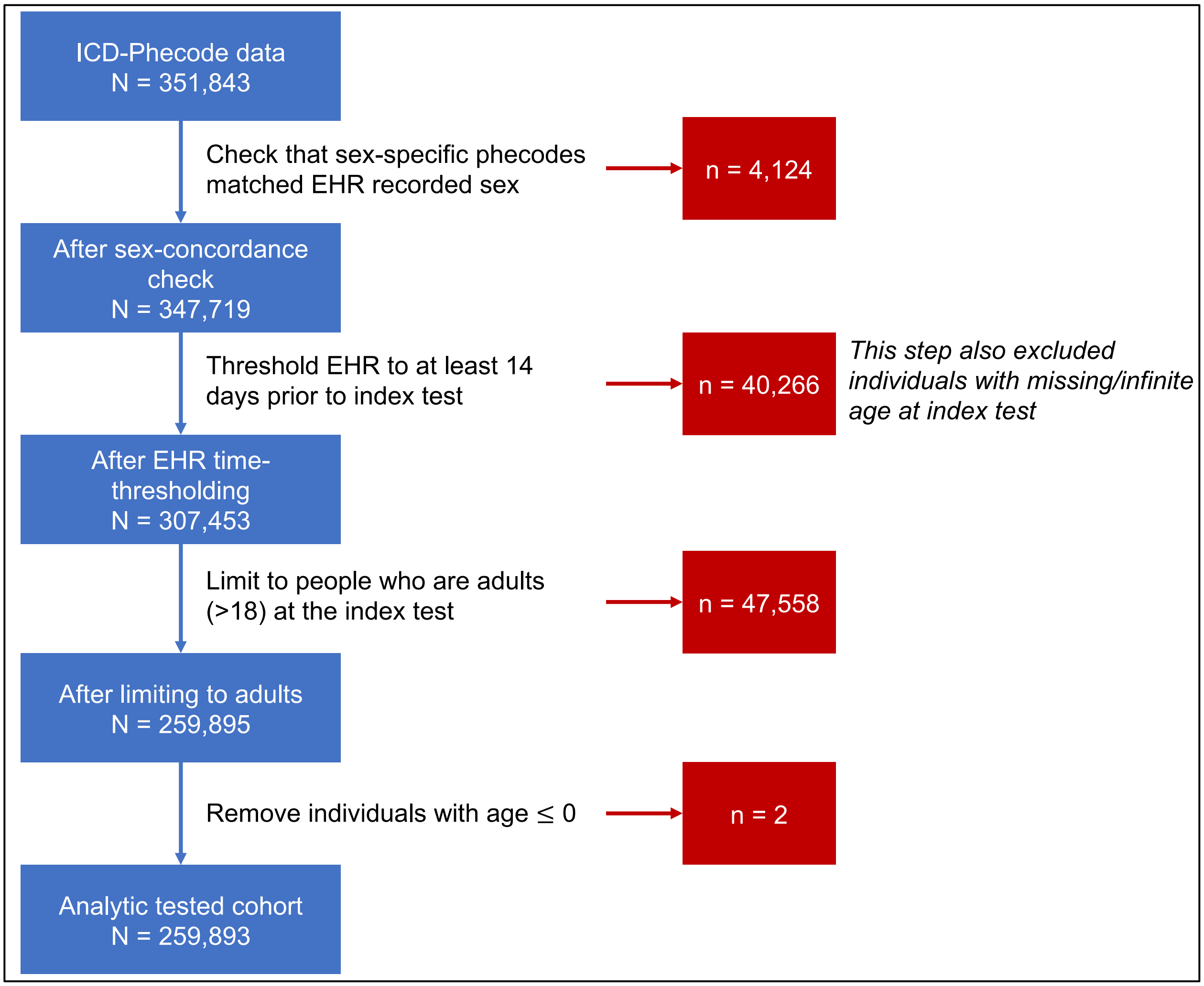

**Figure S2**. An UpSet diagram showing the distribution of cancer treatment patterns among COVID-19 positive individuals with cancer (n = 10,266). Treatment-based analyses were performed using three mutually exclusive categories: chemotherapy (n = 2,449), radiation only (n = 394), and surgery only (n = 111). Treatment was defined according to **Table S2**.

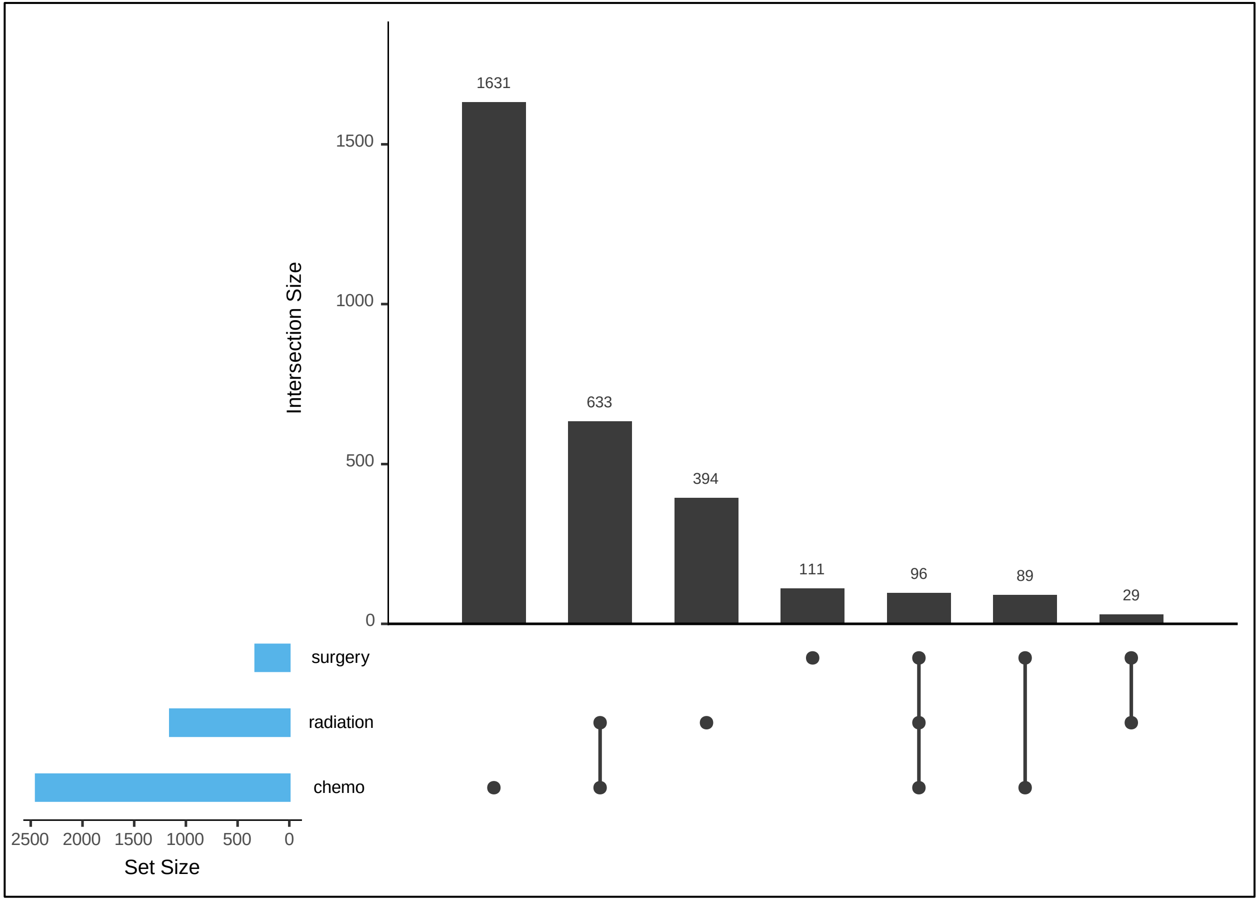

**Table S1**. Phecodes used to identify individuals with cancer, along with their phenotype description and the number of individuals in the tested positive cohort with the corresponding phecode.

| Phecode | Phenotype | Count | Phecode | Phenotype | Count | Phecode | Phenotype | Count |
|---|---|---|---|---|---|---|---|---|
| 145 | Cancer of mouth | 241 | 174 | Breast cancer | 1,120 | 195.3 | Malignant neoplasm of head, face, and neck | 179 |
| 145.1 | Cancer of lip | 16 | 174.1 | Breast cancer [female] | 1,064 | 196 | Radiotherapy | 671 |
| 145.2 | Cancer of tongue | 82 | 174.11 | Malignant neoplasm of female breast | 1,034 | 197 | Chemotherapy | 1,754 |
| 145.3 | Cancer of major salivary glands | 70 | 174.2 | Breast cancer [male] | 8 | 198 | Secondary malignant neoplasm | 1,300 |
| 145.4 | Cancer of the gums | 20 | 174.3 | Neoplasm of uncertain behavior of breast | 76 | 198.1 | Secondary malignancy of lymph nodes | 734 |
| 145.5 | Cancer of the mouth floor | 16 | 175 | Acquired absence of breast | 274 | 198.2 | Secondary malignancy of respiratory organs | 233 |
| 149 | Cancer of larynx, pharynx, nasal cavities | 229 | 180 | Cervical cancer and dysplasia | 1,140 | 198.3 | Secondary malignant neoplasm of digestive systems | 161 |
| 149.1 | Cancer of oropharynx | 72 | 180.1 | Cervical cancer | 170 | 198.4 | Secondary malignant neoplasm of liver | 179 |
| 149.2 | Cancer of nasopharynx | 29 | 180.3 | Cervical intraepithelial neoplasia [CIN] [Cervical dysplasia] | 1,016 | 198.5 | Secondary malignancy of brain/spine | 114 |
| 149.3 | Cancer of hypopharynx | 18 | 182 | Malignant neoplasm of uterus | 218 | 198.6 | Secondary malignancy of bone | 230 |
| 149.4 | Cancer of larynx | 69 | 184 | Cancer of other female genital organs | 377 | 198.7 | Secondary malignant neoplasm of skin | 58 |
| 149.5 | Hx of malignant neoplasm of oral cavity and pharynx | 64 | 184.1 | Malignant neoplasm of ovary and other uterine adnexa | 179 | 199 | Neoplasm of uncertain behavior | 2,304 |
| 149.9 | Cancer of of nasal cavities | 35 | 184.11 | Malignant neoplasm of ovary | 135 | 199.4 | Neurofibromatosis | 50 |
| 150 | Cancer of esophagus | 90 | 184.2 | Cancer of other female genital organs (excluding uterus and ovary) | 169 | 200 | Myeloproliferative disease | 925 |
| 151 | Cancer of stomach | 115 | 185 | Cancer of prostate | 910 | 200.1 | Polycythemia vera | 214 |
| 153 | Colorectal cancer | 563 | 187 | Cancer of other male genital organs | 143 | 201 | Hodgkin's disease | 116 |
| 153.2 | Colon cancer | 443 | 187.1 | Malignant neoplasm of unspecified male genital organ | 24 | 202 | Cancer of other lymphoid, histiocytic tissue | 836 |
| 153.3 | Malignant neoplasm of rectum, rectosigmoid junction, and anus | 296 | 187.2 | Malignant neoplasm of testis | 90 | 202.2 | Non-Hodgkins lymphoma | 745 |
| 155 | Cancer of liver and intrahepatic bile duct | 161 | 187.8 | Neoplasm of uncertain behavior of male genital organs | 39 | 202.21 | Nodular lymphoma | 299 |
| 155.1 | Malignant neoplasm of liver, primary | 128 | 189 | Cancer of urinary organs (incl. kidney and bladder) | 732 | 202.22 | Reticulosarcoma | 137 |
| 157 | Pancreatic cancer | 108 | 189.1 | Cancer of kidney and renal pelvis | 382 | 202.23 | Lymphosarcoma | 21 |
| 158 | Neoplasm of unspecified nature of digestive system | 552 | 189.11 | Malignant neoplasm of kidney, except pelvis | 326 | 202.24 | Large cell lymphoma | 137 |
| 159 | Malignant neoplasm of other and ill-defined sites within the digestive organs and peritoneum | 520 | 189.12 | Malignant neoplasm of renal pelvis | 42 | 204 | Leukemia | 616 |
| 159.2 | Malignant neoplasm of small intestine, including duodenum | 44 | 189.2 | Cancer of bladder | 388 | 204.1 | Lymphoid leukemia | 219 |
| 159.3 | Malignant neoplasm of gallbladder and extrahepatic bile ducts | 27 | 189.21 | Malignant neoplasm of bladder | 350 | 204.11 | Lymphoid leukemia, acute | 97 |
| 159.4 | Malignant neoplasm of retroperitoneum and peritoneum | 54 | 189.4 | Malignant neoplasm of other urinary organs | 93 | 204.12 | Lymphoid leukemia, chronic | 134 |
| 164 | Cancer of intrathoracic organs | 47 | 190 | Cancer of eye | 62 | 204.2 | Myeloid leukemia | 237 |
| 165 | Cancer within the respiratory system | 349 | 191 | Malignant and unknown neoplasms of brain and nervous system | 306 | 204.21 | Myeloid leukemia, acute | 125 |
| 165.1 | Cancer of bronchus; lung | 306 | 191.1 | Cancer of brain and nervous system | 179 | 204.22 | Myeloid leukemia, chronic | 75 |
| 170 | Cancer of bone and connective tissue | 427 | 191.11 | Cancer of brain | 157 | 204.3 | Monocytic leukemia | 34 |
| 170.1 | Bone cancer | 156 | 193 | Thyroid cancer | 322 | 204.4 | Multiple myeloma | 211 |
| 170.2 | Cancer of connective tissue | 324 | 194 | Cancer of other endocrine glands | 202 | 209 | Neuroendocrine tumors | 98 |
| 172.1 | Melanomas of skin, dx or hx | 1,724 | 195 | Cancer, suspected or other | 6,000 | 860 | Bone marrow or stem cell transplant | 227 |
| 172.11 | Melanomas of skin | 751 | 195.1 | Malignant neoplasm, other | 5,878 | | | |

Cells highlighted in pink indicate codes corresponding to lung cancer; cells highlighted in green indicate codes corresponding to melanoma; cells highlighted in blue indicate codes corresponding to breast cancer; cells highlighted in orange indicate codes corresponding to prostate cancer; cells highlighted in yellow indicate codes corresponding to hematologic malignancies

**Table S2**. Codes used to define cancers diagnoses and treatments. Cancer types are highlighted different colors and correspond to the highlighted rows of the same color in Table S1.

| **Definition** | **Code Type** | **Codes** |
|---|---|---|
| Cancer | Phecode | 145, 145.1, 145.2, 145.3, 145.4, 145.5, 149, 149.1, 149.2, 149.3, 149.4, 149.5, 149.9, 150, 151, 153, 153.2, 153.3, 155, 155.1, 157, 158, 159, 159.2, 159.3, 159.4, 164, 165, 165.1, 170, 170.1, 170.2, 172.1, 172.11, 174, 174.1, 174.11, 174.2, 174.3, 175, 180, 180.1, 180.3, 182, 184, 184.1, 184.11, 184.2, 185, 187, 187.1, 187.2, 187.8, 189, 189.1, 189.11, 189.12, 189.2, 189.21, 189.4, 190, 191, 191.1, 191.11, 193, 194, 195, 195.1, 195.3, 196, 197, 198, 198.1, 198.2, 198.3, 198.4, 198.5, 198.6, 198.7, 199, 199.4, 200, 200.1, 201, 202, 202.2, 202.21, 202.22, 202.23, 202.24, 204, 204.1, 204.11, 204.12, 204.2, 204.21, 204.22, 204.3, 204.4, 209, 860 |
| Skin cancer | Phecode | 172.1, 172.11 |
| Hematologic malignancies | Phecode | 200, 200.1, 201, 202, 202.2, 202.21, 202.22, 202.23, 202.24, 204, 204.1, 204.11, 204.12, 204.2, 204.21, 204.22, 204.3, 204.4 |
| Breast cancer | Phecode | 174, 174.1, 174.11, 174.2, 174.3, 175 |
| Prostate cancer | Phecode | 185 |
| Lung cancer | Phecode | 165, 165.1 |
| Chemotherapy | CPT | 96401, 96402, 96403, 96404, 96405, 96406, 96407, 96408, 96409, 96410, 96411, 96412, 96413, 96414, 96415, 96416, 96417, 96420, 96421, 96422, 96423, 96424, 96425<br><br>96360*, 96361*, 96440*, 96441*, 96442*, 96443*, 96444*, 96445*, 96446*, 96447*, 96448*, 96449*, 96450*, 96451*, 96452*, 96453*, 96454*, 96455*, 96456*, 96457*, 96458*, 96459*, 96460*, 96461*, 96462*, 96463*, 96464*, 96465*, 96466*, 96467*, 96468*, 96469*, 96470*, 96471*, 96472*, 96473*, 96474*, 96475*, 96476*, 96477*, 96478*, 96479*, 96480*, 96481*, 96482*, 96483*, 96484*, 96485*, 96486*, 96487*, 96488*, 96489*, 96490*, 96491*, 96492*, 96493*, 96494*, 96495*, 96496*, 96497*, 96498*, 96499*, 96500*, 96501*, 96502*, 96503*, 96504*, 96505*, 96506*, 96507*, 96508*, 96509*, 96510*, 96511*, 96512*, 96513*, 96514*, 96515*, 96516*, 96517*, 96518*, 96519*, 96520*, 96521*, 96522*, 96523*, 96524*, 96525*, 96526*, 96527*, 96528*, 96529*, 96530*, 96531*, 96532*, 96533*, 96534*, 96535*, 96536*, 96537*, 96538*, 96539*, 96540*, 96541*, 96542*, 96543*, 96544*, 96545*, 96546*, 96547*, 96548*, 96549* |

| | | |
|---|---|---|
| | ICD-9 or -10 | 99.25, Z51.1, Z51.11, T80.810, Z92.21, D61.810, D64.81, D70.1, T45.1X5A, V58.11, 999.81, V87.41, 284.11, 285.3, 288.03, E933.1, V66.2 |
| Radiation therapy | CPT | 77295, 77296, 77297, 77298, 77299, 77300, 77301, 77302, 77303, 77304, 77305, 77306, 77307, 77308, 77309, 77310, 77311, 77312, 77313, 77314, 77315, 77316, 77317, 77318, 77319, 77320, 77321, 77322, 77323, 77324, 77325, 77326, 77327, 77328, 77329, 77330, 77331, 77332, 77333, 77334, 77335, 77336, 77337, 77338, 77339, 77340, 77341, 77342, 77343, 77344, 77345, 77346, 77347, 77348, 77349, 77350, 77351, 77352, 77353, 77354, 77355, 77356, 77357, 77358, 77359, 77360, 77361, 77362, 77363, 77364, 77365, 77366, 77367, 77368, 77369, 77370, 77371, 77372, 77373, 77374, 77375, 77376, 77377, 77378, 77379, 77380, 77381, 77382, 77383, 77384, 77385, 77386, 77387, 77399, 77401, 77402, 77403, 77404, 77405, 77406, 77407, 77408, 77409, 77410, 77411, 77412, 77413, 77414, 77415, 77416, 77417, 77418, 77419, 77420, 77421, 77422, 77423, 77424, 77425, 77427, 77428, 77429, 77430, 77431, 77432, 77433, 77434, 77435, 77436, 77437, 77438, 77439, 77440, 77441, 77442, 77443, 77444, 77445, 77446, 77447, 77448, 77449, 77450, 77451, 77452, 77453, 77454, 77455, 77456, 77457, 77458, 77459, 77460, 77461, 77462, 77463, 77464, 77465, 77466, 77467, 77468, 77469, 77470, 77471, 77472, 77473, 77474, 77475, 77476, 77477, 77478, 77479, 77480, 77481, 77482, 77483, 77484, 77485, 77486, 77487, 77488, 77489, 77490, 77491, 77492, 77493, 77494, 77495, 77496, 77497, 77498, 77499, 77520, 77521, 77522, 77523, 77524, 77525, 77600, 77601, 77602, 77603, 77604, 77605, 77606, 77607, 77608, 77609, 77610, 77611, 77612, 77613, 77614, 77615, 77620, 77750, 77751, 77752, 77753, 77754, 77755, 77756, 77757, 77758, 77759, 77760, 77761, 77762, 77763, 77764, 77765, 77766, 77767, 77768, 77769, 77770, 77771, 77772, 77773, 77774, 77775, 77776, 77777, 77778, 77779, 77780, 77781, 77782, 77783, 77784, 77785, 77786, 77787, 77788, 77789, 77790, 77791, 77792, 77793, 77794, 77795, 77796, 77797, 77798, 77799 |
| | ICD-9 or -10 | Z51.0, Z92.3, V58.0, T66.XXX*, 990* |
| Surgery** | CPT | 47120, 47122, 47125, 47130, 47370, 47371, 47379, 47380, 47381, 48120, 48140, 48145, 48146, 48150, 48152, 48153, 48154, 48155, 48160, 19120, 19125, |

| | | |
|---|---|---|
| | | 19126, 19301, 19302, 19303, 19305, 19306, 19307, 44110, 44111, 44140, 44141, 44143, 44144, 44145, 44146, 44147, 44150, 44151, 44155, 44156, 44157, 44158, 44160, 44188, 44204, 44205, 44206, 44207, 44208, 44210, 44211, 44212, 44213, 61518, 61519, 61520, 61521, 61333, 61546, 61548, 61750, 61751, 43610, 43611, 43620, 43621, 43622, 43631, 43632, 43633, 43634, 43117, 43118, 43121, 43122, 43286, 43287, 43288, 43320, 55810, 55812, 55815, 55821, 55831, 55840, 55842, 55845, 55866, 45110, 45111, 45112, 45113, 45114, 45116, 45119, 45126, 45395, 45397, 51550, 51570, 51580, 51555, 51565, 51590, 51575, 51596, 51585, 51595, 51597, 50820 |
| | ICD-9 | 57.6, 57.7, 57.71, 57.79, 1.52, 1.53, 1.59, 85.44, 85.45, 85.46, 85.47, 85.48, 17.31, 17.32, 17.33, 17.34, 17.35, 17.36, 17.39, 45.71, 45.72, 45.73, 45.74, 45.75, 45.76, 45.79, 45.81, 45.82, 45.83, 42.4, 42.41, 42.42, 42.51, 42.52, 42.53, 42.54, 42.55, 42.56, 42.57, 42.58, 42.59, 42.61, 42.62, 42.63, 42.64, 42.65, 42.66, 42.67, 42.68, 42.69, 42.5, 42.6, 43.5, 43.99, 50.22, 50.3, 32.2, 32.29, 32.3, 32.39, 32.41, 32.49, 32.5, 32.59, 32.6, 52.51, 52.52, 52.53, 52.59, 52.6, 52.7, 57.71, 60.4, 60.5, 60.69, 48.5, 48.51, 48.52, 48.59, 48.61, 48.62, 48.63, 48.64, 48.69, 68.8, 43.42, 43.5, 43.6, 43.7, 43.81, 43.82, 43.89, 43.91, 43.99 |
| * Indicates nonspecific codes. Patients with nonspecific treatment codes were assessed as having the treatment only if they had a cancer diagnosis recorded the same day.<br>** All surgery codes were considered nonspecific. | | |

**Table S3**. Phecodes corresponding to the comorbidity classes included in the comorbidity score and their counts in the analytic tested-positive cohort

| Comorbidity class | Phecode | Description | Count |
|---|---|---|---|
| Respiratory | 464 | Acute sinusitis | 5,983 |
| Respiratory | 465 | Acute upper respiratory infections of multiple or unspecified sites | 15,019 |
| Respiratory | 465.2 | Acute pharyngitis | 9,713 |
| Respiratory | 465.4 | Acute laryngitis and tracheitis | 480 |
| Respiratory | 470 | Septal Deviations/Turbinate Hypertrophy | 1,312 |
| Respiratory | 471 | Nasal polyps | 370 |
| Respiratory | 472 | Chronic pharyngitis and nasopharyngitis | 2,092 |
| Respiratory | 473 | Diseases of the larynx and vocal cords | 2,339 |
| Respiratory | 473.1 | Chronic laryngitis | 57 |
| Respiratory | 473.3 | Paralysis/spasm of vocal cords or larynx | 374 |
| Respiratory | 473.4 | Voice disturbance | 1,671 |
| Respiratory | 474 | Acute and chronic tonsillitis | 1,710 |
| Respiratory | 474.1 | Acute tonsillitis | 702 |
| Respiratory | 474.2 | Chronic tonsillitis and adenoiditis | 1,220 |
| Respiratory | 475 | Chronic sinusitis | 4,557 |
| Respiratory | 475.9 | Postnasal drip | 1,341 |
| Respiratory | 476 | Allergic rhinitis | 10,867 |
| Respiratory | 477 | Epistaxis or throat hemorrhage | 1,313 |
| Respiratory | 478 | Throat pain | 678 |
| Respiratory | 479 | Other upper respiratory disease | 6,026 |
| Respiratory | 480 | Pneumonia | 6,215 |
| Respiratory | 480.1 | Bacterial pneumonia | 1,309 |
| Respiratory | 480.11 | Pneumococcal pneumonia | 575 |
| Respiratory | 480.12 | Pseudomonal pneumonia | 133 |
| Respiratory | 480.13 | MRSA pneumonia | 166 |
| Respiratory | 480.2 | Viral pneumonia | 1,146 |
| Respiratory | 480.3 | Pneumonia due to fungus (mycoses) | 186 |
| Respiratory | 480.5 | Bronchopneumonia and lung abscess | 128 |
| Respiratory | 481 | Influenza | 2,070 |
| Respiratory | 483 | Acute bronchitis and bronchiolitis | 4,006 |
| Respiratory | 495 | Asthma | 9,292 |
| Respiratory | 495.1 | Chronic obstructive asthma | 303 |
| Respiratory | 495.11 | Chronic obstructive asthma with exacerbation | 73 |
| Respiratory | 495.2 | Asthma with exacerbation | 2,753 |
| Respiratory | 496 | Chronic airway obstruction | 3,121 |
| Respiratory | 496.1 | Emphysema | 995 |
| Respiratory | 496.2 | Chronic bronchitis | 2,532 |
| Respiratory | 496.21 | Obstructive chronic bronchitis | 2,335 |
| Respiratory | 496.3 | Bronchiectasis | 498 |
| Respiratory | 497 | Bronchitis | 2,237 |
| Respiratory | 498 | Acute bronchospasm | 668 |
| Respiratory | 499 | Cystic fibrosis | 160 |
| Respiratory | 500 | Lung disease due to external agents | 350 |
| Respiratory | 500.1 | Extrinsic allergic alveolitis | 74 |
| Respiratory | 500.2 | Pneumoconiosis | 54 |
| Respiratory | 501 | Pneumonitis due to inhalation of food or vomitus | 515 |
| Respiratory | 502 | Postinflammatory pulmonary fibrosis | 795 |
| Respiratory | 503 | Pulmonary congestion and hypostasis | 529 |
| Respiratory | 504 | Other alveolar and parietoalveolar pneumonopathy | 678 |
| Respiratory | 504.1 | Idiopathic fibrosing alveolitis | 284 |
| Respiratory | 505 | Other pulmonary inflamation or edema | 1,136 |
| Respiratory | 506 | Empyema and pneumothorax | 1,204 |
| Respiratory | 507 | Pleurisy; pleural effusion | 2,119 |
| Respiratory | 508 | Pulmonary collapse; interstitial and compensatory emphysema | 1,769 |
| Respiratory | 509 | Respiratory failure, insufficiency, arrest | 4,046 |
| Respiratory | 509.1 | Respiratory failure | 2,932 |
| Respiratory | 509.2 | Respiratory insufficiency | 1,526 |
| Respiratory | 509.3 | Pulmonary insufficiency or respiratory failure following trauma and surgery | 962 |
| Respiratory | 509.5 | Respiratory arrest | 27 |
| Respiratory | 509.8 | Dependence on respirator [Ventilator] or supplemental oxygen | 1,848 |
| Respiratory | 510 | Other diseases of lung | 3,464 |
| Respiratory | 510.2 | Lung transplant | 185 |
| Respiratory | 512 | Other symptoms of respiratory system | 20,641 |
| Respiratory | 512.1 | Wheezing | 1,834 |
| Respiratory | 512.2 | Painful respiration | 3,013 |
| Respiratory | 512.3 | Abnormal chest sounds | 270 |
| Respiratory | 512.7 | Shortness of breath | 11,738 |
| Respiratory | 512.8 | Cough | 13,143 |
| Respiratory | 512.9 | Other dyspnea | 10,036 |
| Respiratory | 513 | Respiratory abnormalities | 13,826 |
| Respiratory | 513.3 | Hypoventilation | 1,059 |
| Respiratory | 513.31 | Apnea | 535 |
| Respiratory | 513.32 | Orthopnea | 262 |
| Respiratory | 513.4 | Hyperventilation | 426 |
| Respiratory | 513.8 | Disorders of diaphragm | 221 |
| Respiratory | 514 | Abnormal findings examination of lungs | 4,260 |
| Respiratory | 514.1 | Abnormal results of function study of pulmonary system | 327 |
| Respiratory | 514.2 | Solitary pulmonary nodule | 1,826 |
| Respiratory | 516 | Abnormal sputum | 876 |
| Respiratory | 516.1 | Hemoptysis | 705 |
| Respiratory | 519 | Other diseases of respiratory system, not elsewhere classified | 5,115 |
| Respiratory | 519.1 | Tracheostomy complications | 95 |
| Respiratory | 519.2 | Respiratory complications | 415 |
| Respiratory | 519.8 | Other diseases of respiratory system, NEC | 2,282 |
| Respiratory | 519.9 | Symptoms involving respiratory system and other chest symptoms | 2,679 |
| Circulatory | 414 | Other forms of chronic heart disease | 2,474 |
| Circulatory | 414.2 | ASCVD | 716 |

**Table S3**. Phecodes corresponding to the comorbidity classes included in the comorbidity score and their counts in the analytic tested-positive cohort

| Comorbidity class | Phecode | Description | Count |
|---|---|---|---|
| Circulatory | 415 | Pulmonary heart disease | 2,737 |
| Circulatory | 415.1 | Acute pulmonary heart disease | 1,497 |
| Circulatory | 415.11 | Pulmonary embolism and infarction, acute | 1,490 |
| Circulatory | 415.2 | Chronic pulmonary heart disease | 1,513 |
| Circulatory | 415.21 | Primary pulmonary hypertension | 404 |
| Circulatory | 416 | Cardiomegaly | 1,437 |
| Circulatory | 418 | Nonspecific chest pain | 13,438 |
| Circulatory | 418.1 | Precordial pain | 1,200 |
| Circulatory | 420 | Carditis | 1,255 |
| Circulatory | 420.1 | Myocarditis | 120 |
| Circulatory | 420.2 | Pericarditis | 726 |
| Circulatory | 420.21 | Acute pericarditis | 163 |
| Circulatory | 420.22 | Chronic pericarditis | 96 |
| Circulatory | 420.3 | Endocarditis | 522 |
| Circulatory | 425 | Cardiomyopathy | 1,688 |
| Circulatory | 425.1 | Primary/intrinsic cardiomyopathies | 1,594 |
| Circulatory | 425.11 | Hypertrophic obstructive cardiomyopathy | 158 |
| Circulatory | 425.12 | Other hypertrophic cardiomyopathy | 219 |
| Circulatory | 425.2 | Secondary/extrinsic cardiomyopathies | 293 |
| Circulatory | 425.8 | Other cardiomyopathy | 88 |
| Circulatory | 426 | Cardiac conduction disorders | 8,935 |
| Circulatory | 426.2 | Atrioventricular [AV] block | 1,236 |
| Circulatory | 426.21 | First degree AV block | 807 |
| Circulatory | 426.22 | Mobitz II AV block | 46 |
| Circulatory | 426.23 | Second degree AV block | 179 |
| Circulatory | 426.24 | Atrioventricular block, complete | 320 |
| Circulatory | 426.25 | Other heart block | 109 |
| Circulatory | 426.3 | Bundle branch block | 1,619 |
| Circulatory | 426.31 | Right bundle branch block | 1,033 |
| Circulatory | 426.32 | Left bundle branch block | 688 |
| Circulatory | 426.4 | Anomalous atrioventricular excitation | 98 |
| Circulatory | 426.7 | Abnormal electrocardiogram [ECG] [EKG] | 7,821 |
| Circulatory | 426.8 | Other cardiac conduction disorders | 754 |
| Circulatory | 426.9 | Cardiac pacemaker/device in situ | 1,073 |
| Circulatory | 426.91 | Cardiac pacemaker in situ | 665 |
| Circulatory | 426.92 | Cardiac defibrillator in situ | 613 |
| Circulatory | 427 | Cardiac dysrhythmias | 14,422 |
| Circulatory | 427.1 | Paroxysmal tachycardia, unspecified | 2,056 |
| Circulatory | 427.11 | Paroxysmal supraventricular tachycardia | 1,356 |
| Circulatory | 427.12 | Paroxysmal ventricular tachycardia | 999 |
| Circulatory | 427.2 | Atrial fibrillation and flutter | 3,161 |
| Circulatory | 427.21 | Atrial fibrillation | 3,051 |
| Circulatory | 427.22 | Atrial flutter | 1,186 |
| Circulatory | 427.3 | Other specified cardiac dysrhythmias | 6,345 |
| Circulatory | 427.4 | Cardiac arrest and ventricular fibrillation | 439 |
| Circulatory | 427.41 | Ventricular fibrillation and flutter | 189 |
| Circulatory | 427.42 | Cardiac arrest | 319 |
| Circulatory | 427.5 | Arrhythmia (cardiac) NOS | 4,905 |
| Circulatory | 427.6 | Premature beats | 1,912 |
| Circulatory | 427.61 | Supraventricular premature beats | 548 |
| Circulatory | 427.7 | Tachycardia NOS | 5,181 |
| Circulatory | 427.8 | Sinoatrial node dysfunction (Bradycardia) | 565 |
| Circulatory | 427.9 | Palpitations | 5,157 |
| Circulatory | 428 | Congestive heart failure; nonhypertensive | 3,504 |
| Circulatory | 428.1 | Congestive heart failure (CHF) NOS | 2,769 |
| Circulatory | 428.2 | Heart failure NOS | 3,233 |
| Circulatory | 428.3 | Heart failure with reduced EF [Systolic or combined heart failure] | 1,527 |
| Circulatory | 428.4 | Heart failure with preserved EF [Diastolic heart failure] | 1,643 |
| Circulatory | 429 | Ill-defined descriptions and complications of heart disease | 12,866 |
| Circulatory | 429.1 | Heart transplant/surgery | 833 |
| Circulatory | 429.2 | Abnormal function study of cardiovascular system | 6,261 |
| Circulatory | 429.3 | Symptoms involving cardiovascular system | 10,355 |
| Circulatory | 429.9 | Cardiac complications, not elsewhere classified | 331 |
| Circulatory | 430 | Intracranial hemorrhage | 519 |
| Circulatory | 430.1 | Subarachnoid hemorrhage | 181 |
| Circulatory | 430.2 | Intracerebral hemorrhage | 223 |
| Circulatory | 430.3 | Subdural hemorrhage | 183 |
| Circulatory | 433 | Cerebrovascular disease | 3,598 |
| Circulatory | 433.1 | Occlusion and stenosis of precerebral arteries | 1,159 |
| Circulatory | 433.11 | Occlusion of cerebral arteries, with cerebral infarction | 195 |
| Circulatory | 433.12 | Cerebral atherosclerosis | 69 |
| Circulatory | 433.2 | Occlusion of cerebral arteries | 1,856 |
| Circulatory | 433.21 | Cerebral artery occlusion, with cerebral infarction | 1,782 |
| Circulatory | 433.3 | Cerebral ischemia | 1,912 |
| Circulatory | 433.31 | Transient cerebral ischemia | 1,746 |
| Circulatory | 433.32 | Moyamoya disease | 8 |
| Circulatory | 433.5 | Cerebral aneurysm | 309 |
| Circulatory | 433.6 | Acute, but ill-defined cerebrovascular disease | 295 |
| Circulatory | 433.8 | Late effects of cerebrovascular disease | 633 |
| Circulatory | 440 | Atherosclerosis | 1,329 |
| Circulatory | 440.1 | Atherosclerosis of renal artery | 143 |
| Circulatory | 440.2 | Atherosclerosis of the extremities | 736 |
| Circulatory | 440.21 | Atherosclerosis of native arteries of the extremities with ulceration or gangrene | 118 |
| Circulatory | 440.22 | Atherosclerosis of native arteries of the extremities with intermittent claudication | 475 |
| Circulatory | 440.9 | Atherosclerosis of aorta | 437 |
| Circulatory | 441 | Vascular insufficiency of intestine | 320 |
| Circulatory | 441.1 | Acute vascular insufficiency of intestine | 138 |

**Table S3**. Phecodes corresponding to the comorbidity classes included in the comorbidity score and their counts in the analytic tested-positive cohort

| Comorbidity class | Phecode | Description | Count |
|---|---|---|---|
| Circulatory | 441.2 | Chronic vascular insufficiency of intestine | 103 |
| Circulatory | 442 | Other aneurysm | 1,533 |
| Circulatory | 442.1 | Aortic aneurysm | 1,109 |
| Circulatory | 442.11 | Abdominal aortic aneurysm | 545 |
| Circulatory | 442.2 | Aneurysm of iliac artery | 84 |
| Circulatory | 442.3 | Aneurysm of artery of lower extremity | 80 |
| Circulatory | 442.4 | Arterial dissection | 140 |
| Circulatory | 442.8 | Aneurysm of other specified artery | 230 |
| Circulatory | 443 | Peripheral vascular disease | 2,098 |
| Circulatory | 443.1 | Raynaud's syndrome | 686 |
| Circulatory | 443.7 | Peripheral angiopathy in diseases classified elsewhere | 165 |
| Circulatory | 443.8 | Other specified peripheral vascular diseases | 1,855 |
| Circulatory | 443.9 | Peripheral vascular disease, unspecified | 1,378 |
| Circulatory | 444 | Arterial embolism and thrombosis | 528 |
| Circulatory | 444.1 | Arterial embolism and thrombosis of lower extremity artery | 176 |
| Circulatory | 444.2 | Embolism and thrombosis of abdominal aorta | 71 |
| Circulatory | 444.5 | Atheroembolism | 18 |
| Circulatory | 446 | Polyarteritis nodosa and allied conditions | 467 |
| Circulatory | 446.1 | Thromboangiitis obliterans | 9 |
| Circulatory | 446.2 | Acute febrile mucocutaneous lymph node syndrome (Kawasaki disease) | 22 |
| Circulatory | 446.3 | Hypersensitivity angiitis | 55 |
| Circulatory | 446.4 | Wegener's granulomatosis | 90 |
| Circulatory | 446.5 | Giant cell arteritis | 63 |
| Circulatory | 446.6 | Polyarteritis nodosa | 50 |
| Circulatory | 446.7 | Takayasu's disease | 15 |
| Circulatory | 446.8 | Thrombotic microangiopathy | 35 |
| Circulatory | 446.9 | Arteritis NOS | 282 |
| Circulatory | 447 | Other disorders of arteries and arterioles | 1,761 |
| Circulatory | 447.1 | Stricture of artery | 228 |
| Circulatory | 447.7 | Aortic ectasia | 601 |
| Circulatory | 448 | Disease of capillaries | 692 |
| Circulatory | 450 | Noninfectious disorders of lymphatic channels | 952 |
| Circulatory | 451 | Phlebitis and thrombophlebitis | 548 |
| Circulatory | 451.2 | Phlebitis and thrombophlebitis of lower extremities | 173 |
| Circulatory | 452 | Other venous embolism and thrombosis | 3,318 |
| Circulatory | 452.1 | Iatrogenic pulmonary embolism and infarction | 158 |
| Circulatory | 452.2 | Deep vein thrombosis [DVT] | 1,999 |
| Circulatory | 452.8 | Postphlebitic syndrome | 89 |
| Circulatory | 453 | Chronic venous hypertension | 176 |
| Circulatory | 454 | Varicose veins | 2,088 |
| Circulatory | 454.1 | Varicose veins of lower extremity | 1,762 |
| Circulatory | 454.11 | Varicose veins of lower extremity, symptomtic | 1,122 |
| Circulatory | 455 | Hemorrhoids | 5,061 |
| Circulatory | 456 | Chronic venous insufficiency [CVI] | 2,454 |
| Circulatory | 457 | Encounter for long-term (current) use of anticoagulants, antithrombotics, aspirin | 574 |
| Circulatory | 457.2 | Encounter for long-term (current) use of antiplatelets/antithrombotics | 131 |
| Circulatory | 457.3 | Encounter for long-term (current) use of aspirin | 503 |
| Circulatory | 458 | Hypotension | 3,672 |
| Circulatory | 458.1 | Orthostatic hypotension | 1,163 |
| Circulatory | 458.2 | Iatrogenic hypotension | 683 |
| Circulatory | 458.9 | Hypotension NOS | 3,482 |
| Circulatory | 459 | Other disorders of circulatory system | 5,586 |
| Circulatory | 459.1 | Hemorrhage NOS | 599 |
| Circulatory | 459.7 | Blood vessel replaced | 794 |
| Circulatory | 459.9 | Circulatory disease NEC | 4,910 |
| Diabetes | 250.2 | Type 2 diabetes | 7,372 |
| Kidney | 585 | Renal failure | 6,181 |
| Liver | 571 | Chronic liver disease and cirrhosis | 3,670 |
| Autoimmune | 242.1 | Graves' disease | 394 |
| Autoimmune | 250.1 | Type 1 diabetes | 1,435 |
| Autoimmune | 335 | Multiple sclerosis | 349 |
| Autoimmune | 557.1 | Celiac disease | 433 |
| Autoimmune | 694.1 | Vitiligo | 151 |
| Autoimmune | 695.4 | Lupus (localized and systemic) | 783 |
| Autoimmune | 696.4 | Psoriasis | 1,171 |
| Autoimmune | 697 | Sarcoidosis | 418 |
| Autoimmune | 704.1 | Alopecia | 2,056 |
| Autoimmune | 714.1 | Rheumatoid arthritis | 1,448 |
| Autoimmune | 717 | Polymyalgia Rheumatica | 200 |

**Table S4.** Logistic regression odds ratios (95% CI) for COVID-19 outcomes by chemotherapy treatment (without hematologic malignancy patients)

| | Outcomes | | | | n | |
|---|---|---|---|---|---|---|
| | **Severe COVID** | **Hospitalized** | **ICU** | **Deceased** | **Cancer** | **Total** |
| **Any cancer** | | | | | | |
| Unadjusted | **3.57** (3.12, 4.09) | **3.41** (2.95, 3.93) | **2.68** (1.99, 3.61) | **4.79** (3.61, 6.35) | 8,360 | 39,312 |
| Unadjusted (Firth) | **3.58** (3.12, 4.09) | **3.41** (2.95, 3.92) | **2.70** (1.99, 3.60) | **4.82** (3.61, 6.34) | 8,360 | 39,312 |
| Adjustment 1 | **2.26** (1.96, 2.60) | **2.24** (1.93, 2.59) | **1.65** (1.20, 2.21) | **2.27** (1.69, 3.02) | 8,360 | 39,312 |
| Adjustment 2 | **2.27** (1.95, 2.64) | **2.27** (1.94, 2.64) | **1.58** (1.14, 2.16) | **2.20** (1.60, 2.97) | 7,578 | 34,281 |
| Adjustment 3 | **1.76** (1.51, 2.05) | **1.76** (1.50, 2.07) | 1.31 (0.94, 1.79) | **1.66** (1.20, 2.26) | 7,537 | 33,907 |

Adjustment 1: Age, race/ethnicity, sex;
Adjustment 2: Adjustment 1 + Disadvantage Index (quartile)
Adjustment 3: Adjustment 2 + comorbidity score.
Bolded point estimates represents statistical significance at the 95% confidence level.

**Section 1.1. Interaction analysis of cancer diagnosis and other COVID risk factors**

Older age (per 10 years) was associated with higher risk of severe COVID in both cancer [OR (95% CI): 1.29 (1.24, 1.35)] and non-cancer [OR (95% CI): 1.36 (1.32, 1.40)] patients. This association may be stronger for non-cancer patients (p-value for interaction = 0.06). An interaction effect was also found for BMI, with higher BMI increasing odds of severe COVID among those without a cancer diagnosis [OR (95% CI): 1.01 (1.01, 1.02); p-value = 0.01] but a null effect among those with a cancer diagnosis (p-value for interaction <0.01). There was interaction between cancer status and (1) the comorbidity score (OR for severe COVID higher among cancer patients; p-value for interaction = 0.02) and (2) respiratory diseases (OR for severe COVID higher among cancer patients; p-value for interaction = 0.03), and potential interaction with (3) circulatory diseases (OR for severe COVID higher among cancer patients; p-value for interaction = 0.08). The strata-specific odds ratios, their 95% confidence intervals, and the corresponding p-value for interaction for the COVID-19 hospitalization outcome are shown as a forest plot in **Figure S3**. The corresponding plots COVID-19 hospitalization and ICU admission are presented in **Figures S4** and **S5**, which show similar results.

We also identified risk factors that, in conjunction with cancer, were associated with different risk for COVID-19 outcomes in comparison with not having cancer. We found that increasing BMI was associated with increased risk of severe COVID in those without cancer but was not associated in those with cancer. For older patients (≥ 65), having a cancer diagnosis was significantly associated with lower risk of having severe COVID relative to those without cancer. We hypothesize that the behavior of older individuals with cancer diagnoses in their EHR, such as COVID-19 risk mitigation behaviors and increased COVID-19 testing, may play a role in the observed lower severe COVID rates.

**Figure S3**. Severe COVID-19 among cancer (n = 9,333) and non-cancer patients (n = 26,703) who tested positive and had non-missing covariates. The results were from the model logit $P(Y_{SevereCOVID} = 1|X, Cancer, Covariate) = \beta_0 + \beta_X X + \beta_{Cancer} Cancer + \beta_{Int} X \times Cancer + \beta_{Cov} Covariate$, where Covariate = Age + Race/Ethnicity + Sex + Disadvantage Index (quartile) + Comorbidity Score. Reference groups were age: 18 to 35, sex: female, BMI: 18.5 to 25, race: white/non-Hispanic, smoking status: never. A unit change in the continuous age variable is 10 years. The comorbidity score ranges from 0 to 6 and is the sum of indicators for presence of six categories of preexisting conditions in the electronic health record.

| Variable | Cancer OR (95% CI) | No Cancer OR (95% CI) | P−Value Cancer | P−Value No Cancer | P−Value Interaction |
|---|---|---|---|---|---|
| Age | 1.29 (1.24, 1.35) | 1.36 (1.32, 1.40) | <0.01 | <0.01 | 0.06 |
| [35, 50) | 0.55 (0.40, 0.78) | 0.76 (0.62, 0.93) | <0.01 | <0.01 | 0.10 |
| [50,65) | 0.42 (0.30, 0.61) | 0.62 (0.48, 0.82) | <0.01 | <0.01 | 0.02 |
| [65+) | 0.41 (0.27, 0.64) | 0.72 (0.49, 1.04) | <0.01 | 0.08 | <0.01 |
| Female sex | 0.64 (0.56, 0.73) | 0.69 (0.62, 0.76) | <0.01 | <0.01 | 0.41 |
| BMI | 1.00 (0.99, 1.01) | 1.01 (1.01, 1.02) | 0.59 | <0.01 | 0.01 |
| [10, 18.5) | 0.96 (0.45, 1.84) | 2.08 (1.34, 3.11) | 0.91 | <0.01 | 0.05 |
| [25,30) | 0.89 (0.74, 1.07) | 1.05 (0.89, 1.23) | 0.21 | 0.56 | 0.18 |
| [30,200) | 0.84 (0.71, 1.00) | 1.27 (1.10, 1.47) | 0.05 | <0.01 | <0.01 |
| Race/Ethnicity | | | | | |
| African American/Non−Hispanic | 1.88 (1.56, 2.27) | 1.55 (1.34, 1.78) | <0.01 | <0.01 | 0.09 |
| Other/Unknown | 1.02 (0.82, 1.27) | 0.94 (0.80, 1.10) | 0.83 | 0.44 | 0.53 |
| Alcohol consumption | 0.73 (0.63, 0.84) | 0.70 (0.62, 0.79) | <0.01 | <0.01 | 0.66 |
| Smoking status | | | | | |
| Current | 1.19 (1.04, 1.36) | 1.18 (1.05, 1.33) | 0.01 | <0.01 | 0.13 |
| Past | 0.88 (0.63, 1.20) | 1.18 (0.96, 1.43) | 0.43 | 0.12 | 0.96 |
| Population density | 1.04 (0.97, 1.11) | 1.03 (0.97, 1.09) | 0.31 | 0.30 | 0.90 |
| Disadvantage Index (qrtl) | 1.12 (1.05, 1.18) | 1.17 (1.12, 1.23) | <0.01 | <0.01 | 0.19 |
| Affluence Index (qrtl) | 0.95 (0.88, 1.02) | 0.91 (0.85, 0.97) | 0.17 | <0.01 | 0.21 |
| % Hispanic or foreign−born (qrtl) | 1.04 (0.96, 1.14) | 0.97 (0.91, 1.04) | 0.33 | 0.38 | 0.19 |
| % less than high school diploma (qrtl) | 1.10 (1.01, 1.19) | 1.15 (1.07, 1.23) | 0.03 | <0.01 | 0.29 |
| Comorbidity score | 1.44 (1.38, 1.52) | 1.34 (1.29, 1.39) | <0.01 | <0.01 | 0.02 |
| Respiratory diseases | 1.29 (1.07, 1.56) | 1.02 (0.91, 1.14) | <0.01 | 0.77 | 0.03 |
| Circulatory diseases | 1.50 (1.15, 1.98) | 1.16 (1.02, 1.33) | <0.01 | 0.02 | 0.08 |
| Type 2 diabetes | 1.36 (1.19, 1.57) | 1.29 (1.14, 1.46) | <0.01 | <0.01 | 0.54 |
| Kidney diseases | 2.39 (2.08, 2.74) | 2.51 (2.21, 2.85) | <0.01 | <0.01 | 0.59 |
| Liver diseases | 1.30 (1.10, 1.54) | 1.30 (1.09, 1.54) | <0.01 | <0.01 | 1.00 |
| Autoimmune diseases | 1.27 (1.10, 1.47) | 1.09 (0.95, 1.24) | <0.01 | 0.21 | 0.12 |

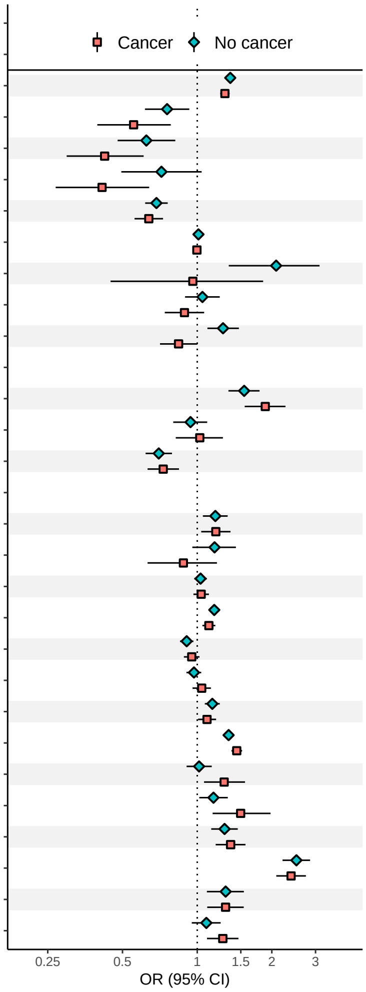

**Figure S4**. COVID-19 hospitalization among cancer (n = 9,333) and non-cancer patients (n = 26,703) who tested positive and had non-missing covariates. The results were from the model $\text{logit } P(Y_{Hospitalization} = 1|X, \text{Cancer}, \text{Covariate}) = \beta_0 + \beta_X X + \beta_{Cancer}\text{CancerStatus} + \beta_{Int} X \times \text{CancerStatus} + \beta_{Cov}\text{Covariate}$, where Covariate = Age + Race/Ethnicity + Sex + Disadvantage Index (quartile) + Comorbidity Score. Reference groups were age: 18 to 35, sex: female, BMI: 18.5 to 25, race: white/non-Hispanic, smoking status: never. A unit change in the continuous age variable is 10 years. The comorbidity score ranges from 0 to 6 and is the sum of indicators for presence of six categories of preexisting conditions in the electronic health record.

| Variable | Cancer OR (95% CI) | No Cancer OR (95% CI) | P−Value Cancer | P−Value No Cancer | P−Value Interaction |
|---|---|---|---|---|---|
| Age | 1.23 (1.18, 1.29) | 1.30 (1.26, 1.34) | <0.01 | <0.01 | 0.05 |
| [35, 50) | 0.54 (0.38, 0.77) | 0.80 (0.65, 0.99) | <0.01 | 0.04 | 0.04 |
| [50,65) | 0.45 (0.31, 0.66) | 0.71 (0.53, 0.93) | <0.01 | 0.01 | <0.01 |
| [65+) | 0.45 (0.29, 0.70) | 0.80 (0.54, 1.19) | <0.01 | 0.27 | <0.01 |
| Female sex | 0.64 (0.56, 0.74) | 0.68 (0.61, 0.76) | <0.01 | <0.01 | 0.53 |
| BMI | 1.00 (0.99, 1.01) | 1.01 (1.00, 1.02) | 0.84 | <0.01 | 0.10 |
| [10, 18.5) | 1.00 (0.45, 1.97) | 2.00 (1.27, 3.02) | 1.00 | <0.01 | 0.10 |
| [25,30) | 0.89 (0.73, 1.08) | 1.07 (0.91, 1.26) | 0.24 | 0.43 | 0.16 |
| [30,200) | 0.91 (0.76, 1.09) | 1.27 (1.10, 1.48) | 0.31 | <0.01 | <0.01 |
| Race/Ethnicity | | | | | |
| African American/Non−Hispanic | 1.96 (1.61, 2.38) | 1.58 (1.36, 1.83) | <0.01 | <0.01 | 0.07 |
| Other/Unknown | 1.02 (0.80, 1.28) | 0.91 (0.76, 1.07) | 0.88 | 0.23 | 0.42 |
| Alcohol consumption | 0.71 (0.61, 0.83) | 0.74 (0.65, 0.84) | <0.01 | <0.01 | 0.69 |
| Smoking status | | | | | |
| Current | 1.15 (0.99, 1.32) | 1.15 (1.02, 1.29) | 0.06 | 0.03 | 0.05 |
| Past | 0.82 (0.58, 1.15) | 1.21 (0.98, 1.48) | 0.26 | 0.08 | 0.99 |
| Population density | 1.09 (1.01, 1.18) | 1.05 (0.99, 1.12) | 0.02 | 0.09 | 0.44 |
| Disadvantage Index (qrtl) | 1.10 (1.03, 1.17) | 1.16 (1.10, 1.22) | <0.01 | <0.01 | 0.21 |
| Affluence Index (qrtl) | 1.01 (0.94, 1.09) | 0.95 (0.89, 1.01) | 0.80 | 0.09 | 0.10 |
| % Hispanic or foreign−born (qrtl) | 1.10 (1.01, 1.20) | 0.98 (0.91, 1.05) | 0.03 | 0.53 | 0.04 |
| % less than high school diploma (qr tl) | 1.07 (0.98, 1.17) | 1.13 (1.05, 1.21) | 0.14 | <0.01 | 0.25 |
| Comorbidity score | 1.47 (1.39, 1.54) | 1.32 (1.26, 1.37) | <0.01 | <0.01 | <0.01 |
| Respiratory diseases | 1.47 (1.19, 1.82) | 1.03 (0.91, 1.16) | <0.01 | 0.66 | <0.01 |
| Circulatory diseases | 1.59 (1.20, 2.14) | 1.13 (0.99, 1.29) | <0.01 | 0.08 | 0.03 |
| Type 2 diabetes | 1.42 (1.22, 1.64) | 1.25 (1.10, 1.43) | <0.01 | <0.01 | 0.20 |
| Kidney diseases | 2.32 (2.01, 2.68) | 2.37 (2.07, 2.71) | <0.01 | <0.01 | 0.81 |
| Liver diseases | 1.38 (1.16, 1.64) | 1.34 (1.12, 1.59) | <0.01 | <0.01 | 0.81 |
| Autoimmune diseases | 1.27 (1.09, 1.48) | 1.02 (0.89, 1.18) | <0.01 | 0.74 | 0.04 |

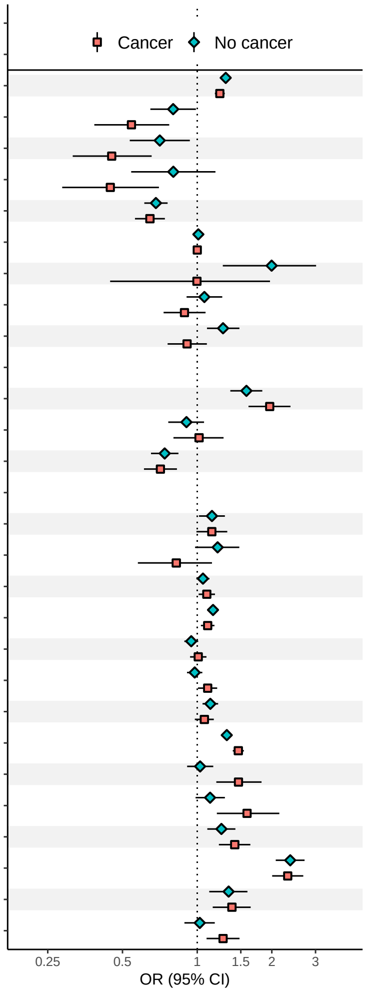

**Figure S5**. COVID-19 ICU admission among cancer (n = 9,333) and non-cancer patients (n = 26,703) who tested positive and had non-missing covariates. The results were from the model logit $P(Y_{ICU} = 1|X, \text{Cancer}, \text{Covariate}) = \beta_0 + \beta_X X + \beta_{Cancer}\text{CancerStatus} + \beta_{Int} X \times \text{CancerStatus} + \beta_{Cov}\text{Covariate}$, where Covariate = Age + Race/Ethnicity + Sex + Disadvantage Index (quartile) + Comorbidity Score. Reference groups were age: 18 to 35, sex: female, BMI: 18.5 to 25, race: white/non-Hispanic, smoking status: never. A unit change in the continuous age variable is 10 years. The comorbidity score ranges from 0 to 6 and is the sum of indicators for presence of six categories of preexisting conditions in the electronic health record.

| Variable | Cancer OR (95% CI) | No Cancer OR (95% CI) | P−Value Cancer | P−Value No Cancer | P−Value Interaction |
|---|---|---|---|---|---|
| Age | 1.26 (1.15, 1.38) | 1.44 (1.35, 1.53) | <0.01 | <0.01 | 0.02 |
| [35, 50) | 0.84 (0.40, 1.90) | 0.96 (0.59, 1.55) | 0.67 | 0.86 | 0.77 |
| [50,65) | 0.87 (0.40, 1.99) | 1.76 (1.01, 3.12) | 0.72 | 0.05 | 0.08 |
| [65+) | 1.05 (0.42, 2.74) | 2.29 (1.06, 4.97) | 0.92 | 0.03 | 0.05 |
| Female sex | 0.54 (0.40, 0.71) | 0.53 (0.42, 0.66) | <0.01 | <0.01 | 0.93 |
| BMI | 1.01 (0.99, 1.03) | 1.01 (0.99, 1.02) | 0.21 | 0.34 | 0.65 |
| [10, 18.5) | 0.85 (0.09, 3.27) | 3.58 (1.72, 6.78) | 0.84 | <0.01 | 0.07 |
| [25,30) | 0.81 (0.55, 1.20) | 0.99 (0.71, 1.38) | 0.29 | 0.95 | 0.44 |
| [30,200) | 1.01 (0.71, 1.46) | 1.20 (0.89, 1.63) | 0.96 | 0.23 | 0.47 |
| Race/Ethnicity | | | | | |
| African American/Non−Hispanic | 2.32 (1.62, 3.29) | 1.84 (1.38, 2.43) | <0.01 | <0.01 | 0.29 |
| Other/Unknown | 1.15 (0.71, 1.77) | 1.21 (0.88, 1.62) | 0.55 | 0.24 | 0.86 |
| Alcohol consumption | 0.83 (0.61, 1.13) | 0.54 (0.42, 0.70) | 0.23 | <0.01 | 0.03 |
| Smoking status | | | | | |
| Current | 1.33 (1.00, 1.77) | 1.07 (0.84, 1.35) | 0.05 | 0.59 | 0.73 |
| Past | 0.64 (0.26, 1.34) | 0.76 (0.45, 1.20) | 0.26 | 0.25 | 0.23 |
| Population density | 1.08 (0.93, 1.26) | 1.03 (0.91, 1.16) | 0.29 | 0.66 | 0.57 |
| Disadvantage Index (qrtl) | 1.15 (1.02, 1.30) | 1.17 (1.06, 1.29) | 0.03 | <0.01 | 0.83 |
| Affluence Index (qrtl) | 0.99 (0.85, 1.14) | 0.95 (0.84, 1.08) | 0.85 | 0.44 | 0.64 |
| % Hispanic or foreign−born (qrtl) | 1.00 (0.83, 1.19) | 0.99 (0.86, 1.14) | 0.97 | 0.88 | 0.95 |
| % less than high school diploma (qr tl) | 1.05 (0.88, 1.24) | 1.10 (0.96, 1.27) | 0.60 | 0.18 | 0.59 |
| Comorbidity score | 1.37 (1.24, 1.52) | 1.25 (1.15, 1.35) | <0.01 | <0.01 | 0.13 |
| Respiratory diseases | 1.29 (0.88, 1.97) | 0.98 (0.77, 1.24) | 0.20 | 0.84 | 0.22 |
| Circulatory diseases | 1.26 (0.74, 2.34) | 0.99 (0.76, 1.31) | 0.41 | 0.95 | 0.43 |
| Type 2 diabetes | 1.73 (1.30, 2.29) | 1.44 (1.13, 1.84) | <0.01 | <0.01 | 0.33 |
| Kidney diseases | 1.90 (1.43, 2.53) | 2.36 (1.83, 3.04) | <0.01 | <0.01 | 0.24 |
| Liver diseases | 1.39 (0.97, 1.93) | 0.98 (0.66, 1.40) | 0.07 | 0.91 | 0.17 |
| Autoimmune diseases | 1.07 (0.78, 1.46) | 0.88 (0.65, 1.16) | 0.65 | 0.37 | 0.34 |

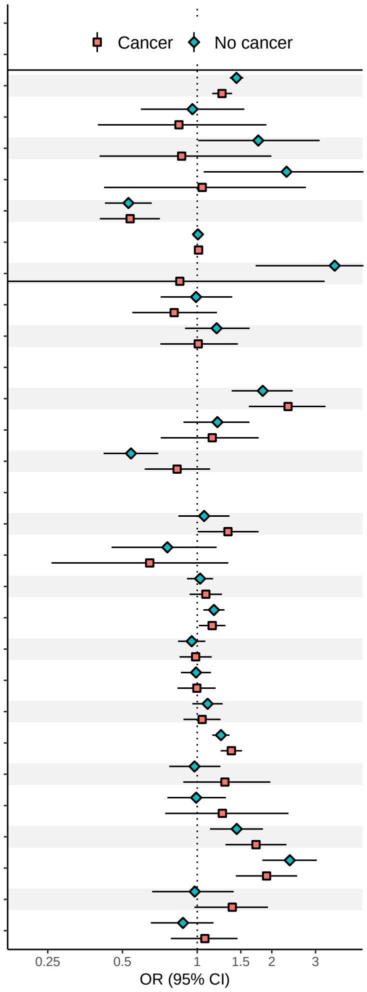

**Section 2**. Cancer recency analysis results

**Table S5**. Logistic regression odds ratios (95% CI) for COVID-19 outcomes by cancer treatment type using restricted electronic health record for cancer recency analysis.

| | Outcomes | | | | n | |
|---|---|---|---|---|---|---|
| | Severe COVID | Hospitalized | ICU | Deceased | Cancer/ treatment group | Total |
| **Chemotherapy** | | | | | | |
| Unadjusted | **4.44** (3.82, 5.15) | **4.08** (3.49, 4.77) | **3.55** (2.60, 4.83) | **6.45** (4.82, 8.63) | 1,158 | 41,218 |
| Unadjusted (Firth) | **4.44** (3.82, 5.15) | **4.09** (3.48, 4.77) | **3.58** (2.60, 4.82) | **6.49** (4.81, 8.61) | 1,158 | 41,218 |
| Adjustment 1 | **2.88** (2.46, 3.35) | **2.74** (2.32, 3.22) | **2.20** (1.58, 2.98) | **3.14** (2.31, 4.20) | 1,158 | 41,218 |
| Adjustment 2 | **2.94** (2.49, 3.45) | **2.84** (2.39, 3.36) | **2.15** (1.53, 2.96) | **2.91** (2.09, 3.99) | 1,014 | 36,036 |
| Adjustment 3 | **2.41** (2.04, 2.84) | **2.33** (1.96, 2.77) | **1.81** (1.28, 2.50) | **2.33** (1.66, 3.20) | 1,014 | 36,036 |
| **Radiation only** | | | | | | |
| Unadjusted | **4.00** (2.81, 5.70) | **3.86** (2.67, 5.58) | **5.79** (3.27, 10.26) | **7.08** (3.81, 13.17) | 201 | 41,218 |
| Unadjusted (Firth) | **4.04** (2.81, 5.68) | **3.90** (2.67, 5.55) | **5.99** (3.26, 10.12) | **7.37** (3.79, 12.94) | 201 | 41,218 |
| Adjustment 1 | **2.30** (1.59, 3.26) | **2.34** (1.58, 3.35) | **3.30** (1.78, 5.64) | **3.11** (1.59, 5.55) | 201 | 41,218 |
| Adjustment 2 | **2.03** (1.33, 2.99) | **2.16** (1.40, 3.21) | **3.15** (1.61, 5.59) | **2.37** (1.02, 4.72) | 176 | 36,036 |
| Adjustment 3 | **1.74** (1.14, 2.59) | **1.86** (1.20, 2.79) | **2.77** (1.41, 4.93) | 1.91 (0.82, 3.84) | 176 | 36,036 |
| **Surgery only** | | | | | | |
| Unadjusted | 2.52 (0.98, 6.46) | **2.77** (1.08, 7.12) | 2.26 (0.31, 16.55) | - | 38 | 41,218 |
| Unadjusted (Firth) | 2.73 (0.98, 6.21) | **3.01** (1.08, 6.84) | 3.35 (0.38, 12.55) | 1.59 (0.01, 11.17) | 38 | 41,218 |
| Adjustment 1 | 1.70 (0.60, 3.97) | 1.96 (0.70, 4.58) | 2.04 (0.23, 7.87) | 0.73 (0.01, 5.40) | 38 | 41,218 |
| Adjustment 2 | 1.66 (0.59, 3.92) | 1.92 (0.68, 4.52) | 1.83 (0.20, 7.13) | 0.68 (0.01, 5.05) | 37 | 36,036 |
| Adjustment 3 | 1.22 (0.42, 2.95) | 1.43 (0.50, 3.43) | 1.39 (0.15, 5.48) | 0.45 (0.00, 3.44) | 37 | 35,036 |

Adjustment 1: Age, race/ethnicity, sex;
Adjustment 2: Adjustment 1 + Disadvantage Index (quartile)
Adjustment 3: Adjustment 2 + comorbidity score.
Bolded point estimates represents statistical significance at the 95% confidence level.

**Figure S6.** Severe COVID-19 among recent cancer (within three years of positive COVID-19 test; n = 4,260) and non-cancer patients (n = 31,776) who tested positive and have non-missing covariates. The results were from the model logit $P(Y_{SevereCOVID} = 1|X, \text{Cancer}, \text{Covariate}) = \beta_0 + \beta_X X + \beta_{Cancer}\text{Cancer} + \beta_{Int} X \times \text{Cancer} + \beta_{Cov}\text{Covariate}$, where Covariate = Age + Race/Ethnicity + Sex + Disadvantage Index (quartile) + Comorbidity Score. Reference groups were age: 18 to 35, sex: female, BMI: 18.5 to 25, race: white/non-Hispanic, smoking status: never. A unit change in the continuous age variable is 10 years. The comorbidity score ranges from 0 to 6 and is the sum of indicators for presence of six categories of preexisting conditions in the electronic health record.

| Variable | Cancer OR (95% CI) | No Cancer OR (95% CI) | P−Value Cancer | P−Value No Cancer | P−Value Interaction |
|---|---|---|---|---|---|
| Age | 1.19 (1.12, 1.27) | 1.36 (1.32, 1.40) | <0.01 | <0.01 | <0.01 |
| [35, 50) | 0.55 (0.35, 0.87) | 0.76 (0.62, 0.93) | 0.01 | <0.01 | 0.19 |
| [50,65) | 0.36 (0.23, 0.57) | 0.62 (0.48, 0.82) | <0.01 | <0.01 | 0.01 |
| [65+) | 0.31 (0.19, 0.52) | 0.72 (0.50, 1.04) | <0.01 | 0.08 | <0.01 |
| Female sex | 0.72 (0.60, 0.87) | 0.68 (0.62, 0.76) | <0.01 | <0.01 | 0.65 |
| BMI | 0.99 (0.98, 1.01) | 1.01 (1.01, 1.02) | 0.23 | <0.01 | <0.01 |
| [10, 18.5) | 1.01 (0.38, 2.26) | 2.08 (1.34, 3.11) | 0.99 | <0.01 | 0.13 |
| [25,30) | 0.76 (0.59, 0.98) | 1.05 (0.90, 1.24) | 0.03 | 0.54 | 0.03 |
| [30,200) | 0.77 (0.61, 0.98) | 1.27 (1.10, 1.47) | 0.03 | <0.01 | <0.01 |
| Race/Ethnicity | | | | | |
| African American/Non−Hispanic | 1.53 (1.15, 2.01) | 1.55 (1.34, 1.79) | <0.01 | <0.01 | 0.93 |
| Other/Unknown | 0.87 (0.63, 1.19) | 0.94 (0.80, 1.10) | 0.39 | 0.45 | 0.67 |
| Alcohol consumption | 0.66 (0.53, 0.82) | 0.70 (0.62, 0.79) | <0.01 | <0.01 | 0.62 |
| Smoking status | | | | | |
| Current | 1.11 (0.92, 1.34) | 1.19 (1.06, 1.33) | 0.28 | <0.01 | 0.28 |
| Past | 0.90 (0.57, 1.38) | 1.18 (0.96, 1.44) | 0.65 | 0.12 | 0.55 |
| Population density | 0.96 (0.87, 1.06) | 1.03 (0.97, 1.10) | 0.42 | 0.27 | 0.20 |
| Disadvantage Index (qrtl) | 1.05 (0.96, 1.14) | 1.17 (1.12, 1.23) | 0.30 | <0.01 | 0.02 |
| Affluence Index (qrtl) | 0.97 (0.88, 1.06) | 0.91 (0.85, 0.97) | 0.47 | <0.01 | 0.19 |
| % Hispanic or foreign−born (qrtl) | 1.00 (0.89, 1.13) | 0.97 (0.91, 1.04) | 0.95 | 0.38 | 0.63 |
| % less than high school diploma (qr tl) | 1.04 (0.94, 1.16) | 1.15 (1.07, 1.23) | 0.44 | <0.01 | 0.09 |
| Comorbidity score | 1.43 (1.33, 1.53) | 1.34 (1.29, 1.39) | <0.01 | <0.01 | 0.12 |
| Respiratory diseases | 1.46 (1.15, 1.86) | 1.01 (0.90, 1.14) | <0.01 | 0.80 | <0.01 |
| Circulatory diseases | 1.76 (1.24, 2.57) | 1.16 (1.02, 1.33) | <0.01 | 0.03 | 0.03 |
| Type 2 diabetes | 1.22 (1.00, 1.49) | 1.29 (1.14, 1.46) | 0.04 | <0.01 | 0.65 |
| Kidney diseases | 2.10 (1.73, 2.54) | 2.50 (2.20, 2.84) | <0.01 | <0.01 | 0.12 |
| Liver diseases | 1.40 (1.09, 1.77) | 1.30 (1.09, 1.54) | <0.01 | <0.01 | 0.63 |
| Autoimmune diseases | 1.20 (0.97, 1.49) | 1.09 (0.95, 1.24) | 0.09 | 0.23 | 0.42 |

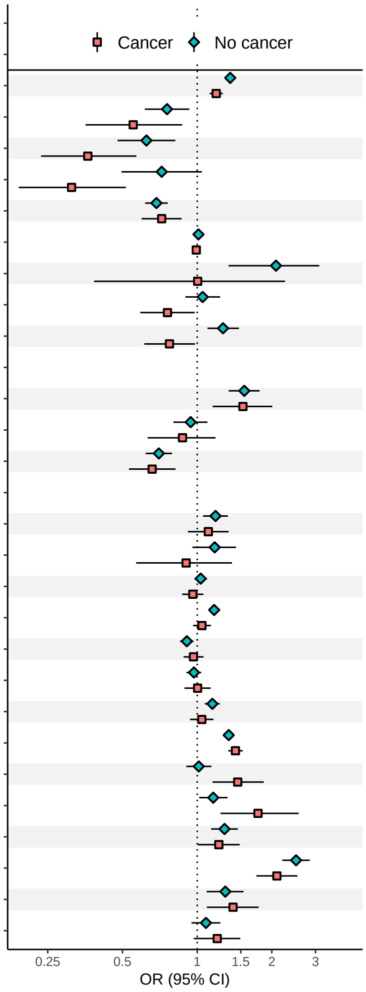

**Figure S7.** COVID-19 hospitalization among recent cancer (within three years of positive COVID-19 test; n = 4,260) and non-cancer patients (n = 31,776) who tested positive and have non-missing covariates. The results were from the model logit $P(Y_{Hospitalization} = 1|X, Cancer, Covariate) = \beta_0 + \beta_X X + \beta_{Cancer} Cancer + \beta_{Int} X \times Cancer + \beta_{Cov} Covariate$, where Covariate = Age + Race/Ethnicity + Sex + Disadvantage Index (quartile) + Comorbidity Score. Reference groups were age: 18 to 35, sex: female, BMI: 18.5 to 25, race: white/non-Hispanic, smoking status: never. A unit change in the continuous age variable is 10 years. The comorbidity score ranges from 0 to 6 and is the sum of indicators for presence of six categories of preexisting conditions in the electronic health record.

| Variable | Cancer OR (95% CI) | No Cancer OR (95% CI) | P−Value Cancer | P−Value No Cancer | P−Value Interaction |
|---|---|---|---|---|---|
| Age | 1.13 (1.06, 1.21) | 1.30 (1.26, 1.34) | <0.01 | <0.01 | <0.01 |
| [35, 50) | 0.53 (0.34, 0.84) | 0.80 (0.65, 0.99) | <0.01 | 0.04 | 0.09 |
| [50,65) | 0.38 (0.24, 0.60) | 0.71 (0.54, 0.94) | <0.01 | 0.02 | <0.01 |
| [65+) | 0.33 (0.20, 0.56) | 0.81 (0.54, 1.19) | <0.01 | 0.28 | <0.01 |
| Female sex | 0.74 (0.61, 0.90) | 0.68 (0.61, 0.76) | <0.01 | <0.01 | 0.48 |
| BMI | 1.00 (0.98, 1.01) | 1.01 (1.00, 1.02) | 0.54 | <0.01 | 0.06 |
| [10, 18.5) | 0.99 (0.35, 2.32) | 1.99 (1.27, 3.01) | 0.98 | <0.01 | 0.16 |
| [25,30) | 0.75 (0.57, 0.98) | 1.07 (0.91, 1.27) | 0.03 | 0.41 | 0.02 |
| [30,200) | 0.83 (0.65, 1.06) | 1.27 (1.10, 1.48) | 0.14 | <0.01 | <0.01 |
| Race/Ethnicity | | | | | |
| African American/Non−Hispanic | 1.67 (1.25, 2.21) | 1.58 (1.36, 1.83) | <0.01 | <0.01 | 0.73 |
| Other/Unknown | 0.77 (0.54, 1.08) | 0.91 (0.77, 1.07) | 0.14 | 0.24 | 0.41 |
| Alcohol consumption | 0.62 (0.50, 0.78) | 0.74 (0.65, 0.84) | <0.01 | <0.01 | 0.19 |
| Smoking status | | | | | |
| Current | 1.07 (0.88, 1.31) | 1.15 (1.02, 1.30) | 0.51 | 0.02 | 0.17 |
| Past | 0.85 (0.51, 1.34) | 1.21 (0.98, 1.48) | 0.49 | 0.07 | 0.55 |
| Population density | 1.02 (0.92, 1.14) | 1.06 (0.99, 1.12) | 0.66 | 0.08 | 0.60 |
| Disadvantage Index (qrtl) | 1.02 (0.94, 1.12) | 1.16 (1.10, 1.22) | 0.59 | <0.01 | 0.02 |
| Affluence Index (qrtl) | 1.04 (0.94, 1.14) | 0.95 (0.89, 1.01) | 0.47 | 0.10 | 0.07 |
| % Hispanic or foreign−born (qrtl) | 1.06 (0.94, 1.21) | 0.98 (0.91, 1.05) | 0.34 | 0.53 | 0.25 |
| % less than high school diploma (qrtl) | 1.01 (0.90, 1.14) | 1.13 (1.05, 1.22) | 0.82 | <0.01 | 0.08 |
| Comorbidity score | 1.45 (1.35, 1.56) | 1.32 (1.26, 1.37) | <0.01 | <0.01 | 0.02 |
| Respiratory diseases | 1.70 (1.31, 2.22) | 1.03 (0.91, 1.16) | <0.01 | 0.69 | <0.01 |
| Circulatory diseases | 1.87 (1.28, 2.83) | 1.13 (0.98, 1.29) | <0.01 | 0.08 | 0.01 |
| Type 2 diabetes | 1.26 (1.02, 1.55) | 1.25 (1.10, 1.43) | 0.03 | <0.01 | 0.97 |
| Kidney diseases | 2.06 (1.68, 2.52) | 2.37 (2.07, 2.70) | <0.01 | <0.01 | 0.25 |
| Liver diseases | 1.52 (1.18, 1.94) | 1.34 (1.12, 1.59) | <0.01 | <0.01 | 0.40 |
| Autoimmune diseases | 1.20 (0.95, 1.50) | 1.02 (0.89, 1.17) | 0.13 | 0.77 | 0.24 |

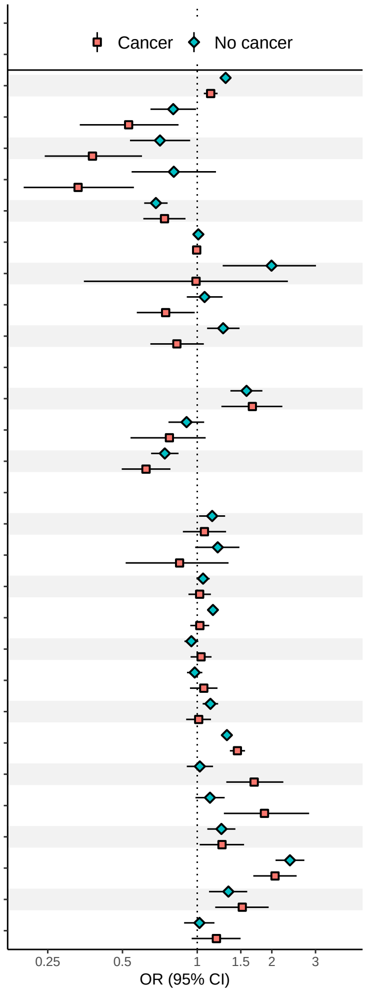

**Figure S8.** COVID-19 ICU admission among recent cancer (within three years of positive COVID-19 test; n = 4,260) and non-cancer patients (n = 31,776) who tested positive and have non-missing covariates. The results were from the model logit $P(Y_{ICU} = 1|X, Cancer, Covariate) = \beta_0 + \beta_X X + \beta_{Cancer} Cancer + \beta_{Int} X \times Cancer + \beta_{Cov} Covariate$, where Covariate = Age + Race/Ethnicity + Sex + Disadvantage Index (quartile) + Comorbidity Score. Reference groups were age: 18 to 35, sex: female, BMI: 18.5 to 25, race: white/non-Hispanic, smoking status: never. A unit change in the continuous age variable is 10 years. The comorbidity score ranges from 0 to 6 and is the sum of indicators for presence of six categories of preexisting conditions in the electronic health record.

| Variable | Cancer OR (95% CI) | No Cancer OR (95% CI) | P−Value Cancer | P−Value No Cancer | P−Value Interaction |
|---|---|---|---|---|---|
| Age | 1.13 (1.00, 1.29) | 1.44 (1.35, 1.53) | 0.05 | <0.01 | <0.01 |
| [35, 50) | 0.57 (0.23, 1.49) | 0.95 (0.59, 1.54) | 0.24 | 0.85 | 0.32 |
| [50,65) | 0.47 (0.20, 1.20) | 1.75 (1.00, 3.10) | 0.11 | 0.05 | <0.01 |
| [65+) | 0.57 (0.22, 1.61) | 2.27 (1.05, 4.93) | 0.28 | 0.04 | <0.01 |
| Female sex | 0.60 (0.40, 0.88) | 0.53 (0.42, 0.66) | <0.01 | <0.01 | 0.58 |
| BMI | 1.00 (0.97, 1.03) | 1.01 (0.99, 1.02) | 0.99 | 0.35 | 0.67 |
| [10, 18.5) | 0.38 (0.00, 2.82) | 3.59 (1.72, 6.79) | 0.43 | <0.01 | 0.04 |
| [25,30) | 0.71 (0.43, 1.19) | 0.99 (0.71, 1.38) | 0.19 | 0.95 | 0.29 |
| [30,200) | 0.76 (0.48, 1.23) | 1.20 (0.89, 1.63) | 0.26 | 0.24 | 0.11 |
| Race/Ethnicity | | | | | |
| African American/Non−Hispanic | 2.04 (1.21, 3.31) | 1.84 (1.38, 2.43) | <0.01 | <0.01 | 0.72 |
| Other/Unknown | 0.94 (0.46, 1.73) | 1.21 (0.88, 1.63) | 0.85 | 0.24 | 0.49 |
| Alcohol consumption | 0.79 (0.51, 1.23) | 0.54 (0.42, 0.70) | 0.29 | <0.01 | 0.14 |
| Smoking status | | | | | |
| Current | 1.42 (0.97, 2.11) | 1.07 (0.84, 1.35) | 0.07 | 0.59 | 0.21 |
| Past | 0.29 (0.03, 1.10) | 0.76 (0.45, 1.20) | 0.07 | 0.25 | 0.21 |
| Population density | 0.99 (0.81, 1.21) | 1.03 (0.91, 1.16) | 0.90 | 0.64 | 0.72 |
| Disadvantage Index (qrtl) | 1.01 (0.85, 1.20) | 1.17 (1.06, 1.29) | 0.91 | <0.01 | 0.13 |
| Affluence Index (qrtl) | 1.00 (0.83, 1.21) | 0.95 (0.84, 1.08) | 0.99 | 0.44 | 0.61 |
| % Hispanic or foreign−born (qrtl) | 0.97 (0.75, 1.23) | 0.99 (0.86, 1.14) | 0.79 | 0.88 | 0.87 |
| % less than high school diploma (qrtl) | 1.02 (0.82, 1.27) | 1.10 (0.96, 1.27) | 0.85 | 0.18 | 0.52 |
| Comorbidity score | 1.41 (1.23, 1.61) | 1.25 (1.15, 1.35) | <0.01 | <0.01 | 0.13 |
| Respiratory diseases | 1.66 (1.01, 2.89) | 0.97 (0.77, 1.24) | 0.05 | 0.82 | 0.05 |
| Circulatory diseases | 1.25 (0.65, 2.75) | 0.99 (0.76, 1.30) | 0.53 | 0.93 | 0.53 |
| Type 2 diabetes | 1.55 (1.05, 2.29) | 1.44 (1.12, 1.84) | 0.03 | <0.01 | 0.74 |
| Kidney diseases | 1.87 (1.27, 2.75) | 2.35 (1.82, 3.03) | <0.01 | <0.01 | 0.31 |
| Liver diseases | 1.62 (1.00, 2.52) | 0.98 (0.66, 1.40) | 0.05 | 0.90 | 0.10 |
| Autoimmune diseases | 1.15 (0.73, 1.77) | 0.87 (0.65, 1.16) | 0.53 | 0.36 | 0.30 |

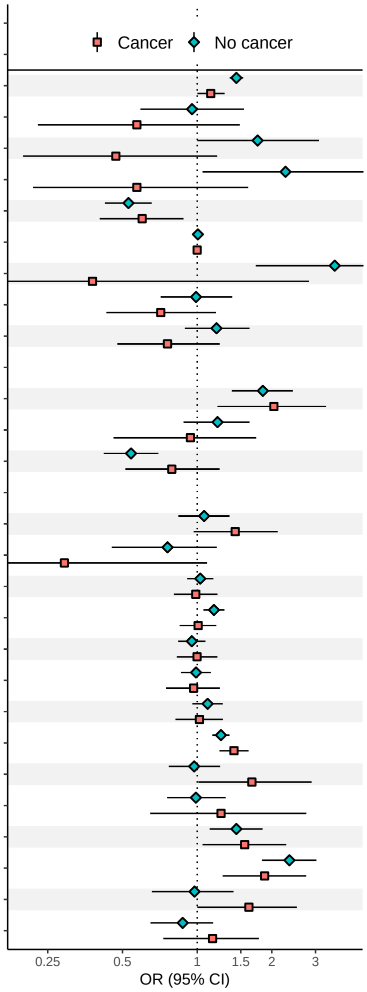
